# The burden of tuberculosis among foreign-born Canadians: estimates with dynamic models

**DOI:** 10.1101/2025.01.23.24318580

**Authors:** Jeremy Chiu, William Ruth, Alexander Rutherford, Kezia Wijaya, Timothy Lee, JF Williams, Albert Wong

## Abstract

**Background:** Despite only comprising about a quarter of the total population of Canada, foreign-born individuals bear about three-quarters of the burden of active tuberculosis (TB) cases. New immigrants arriving in Canada are screened for active TB, but generally not for latent TB infection (LTBI); thus the burden of LTBI among foreign-born Canadians is not well understood.

**Methods:** To investigate the impact of immigration on the burden of TB among foreign-born Canadians, we develop an SEIR-compartment model that distinguishes between actively infected, latently infected, and uninfected individuals. Unknown parameters are calibrated to reports on the incidence and prevalence of active TB in Canada. We validate our model by comparing model computed quantities to other estimates of tuberculosis burden among foreign-born Canadians, including an estimate of the prevalence of LTBI among immigrants entering Canada.

**Results:** If the profile and number of immigrants arriving into Canada in the next decade is similar to the past decade, our model predicts that among the foreign-born population, Canada will not meet the End TB 2035 goal of reducing incidence by 90% compared to 2015. In fact, Canada would still fail to meet the incidence goal if no new immigrants are allowed to enter, primarily due to the activation of foreign-born Canadians with LTBI.

**Author summary:** Our model examines how immigration affects the burden of tuberculosis among foreign-born Canadians. We fit a compartment model to calibration data, then find a feasible parameter set based on validation data. We forecast the incidence of TB in 2035 and demonstrate that regardless of which WHO geographic region immigrants originate from, Canada will fail to meet the WHO End TB’s 2035 incidence goal (90% less than 2015) among the foreign-born population.

## Introduction

Every two minutes, about 5 people die of tuberculosis (TB): the World Health Organization (WHO) estimated that in 2022, TB caused 1.30 million deaths, making it the second deadliest infectious pathogen (second only to COVID-19) (1). An additional 10.6 million developed active TB in 2022, which is more than the estimates from 2020 and 2021. In the absence of HIV, the 10-year case fatality rate of active TB is reported to be 53-86% (weighted average 70%) (2).

### Epidemiology of tuberculosis in Canada

The extent of TB is well-documented in Canada, including reports (3; 4), data about the burden of TB in Canada (5), and reports specific to the foreign-born population (6). Additionally, tuberculosis inequitably affects the Canadian Indigenous People: in 2017, 71% of cases among Canadian-born citizens came from people identifying as Indigenous (7). Outbreaks in Indigenous communities has been reported (8) and modeled (9; 10).

In Canada, TB disproportionately affects foreign-born individuals. In 2021, although only about 23% of individuals residing in Canada were foreign-born (11), about 75% of confirmed active TB cases were from individuals born outside of Canada (5). There have been various models used to estimate the burden of TB among foreign-born Canadians (12; 13; 14). More generally, numerous authors have studied the burden of TB involving immigration from a high-incidence to low-incidence setting, including in the United States (15), the United Kingdom (16), and the Netherlands (17). A recent study investigated the cost-effectiveness of various LTBI screening strategies in the United States (18).

### Our work

In this paper, we develop a compartmental model to study the dynamics of tuberculosis (TB) among foreign-born Canadians, incorporating infection, progression between latent and active TB, and the effects of immigration.” We then train and validate the model using incidence and prevalence reports (5; 6). Finally, we develop methods to forecast the immigration and TB burden in Canada, and predict that among the foreign-born population, Canada will not achieve the *The End TB Strategy* 2035 incidence goals.

Similar to the work of Jordan et al. (12), we estimate the prevalence of tuberculosis infection among the foreign-born Canadian population. Although we used different methodology, our estimates of the prevalence of tuberculosis infection are very similar. Our model additionally estimates the prevalence of active TB, the prevalence of recent infection, and the rates of TB transmission and activation.

## Methods

### Estimating the burden of TB among new Canadian immigrants

To estimate the burden of LTBI among new immigrants, we take a weighted average of two quantities: the number of immigrants from each country that are entering Canada (StatCan’s reported immigration data (19)), and the prevalence of LTBI in each of those countries (Houben and Dodd (2016) (20)). A snapshot of these 2 data sets is visualized in Fig 1.

**Fig 1.**
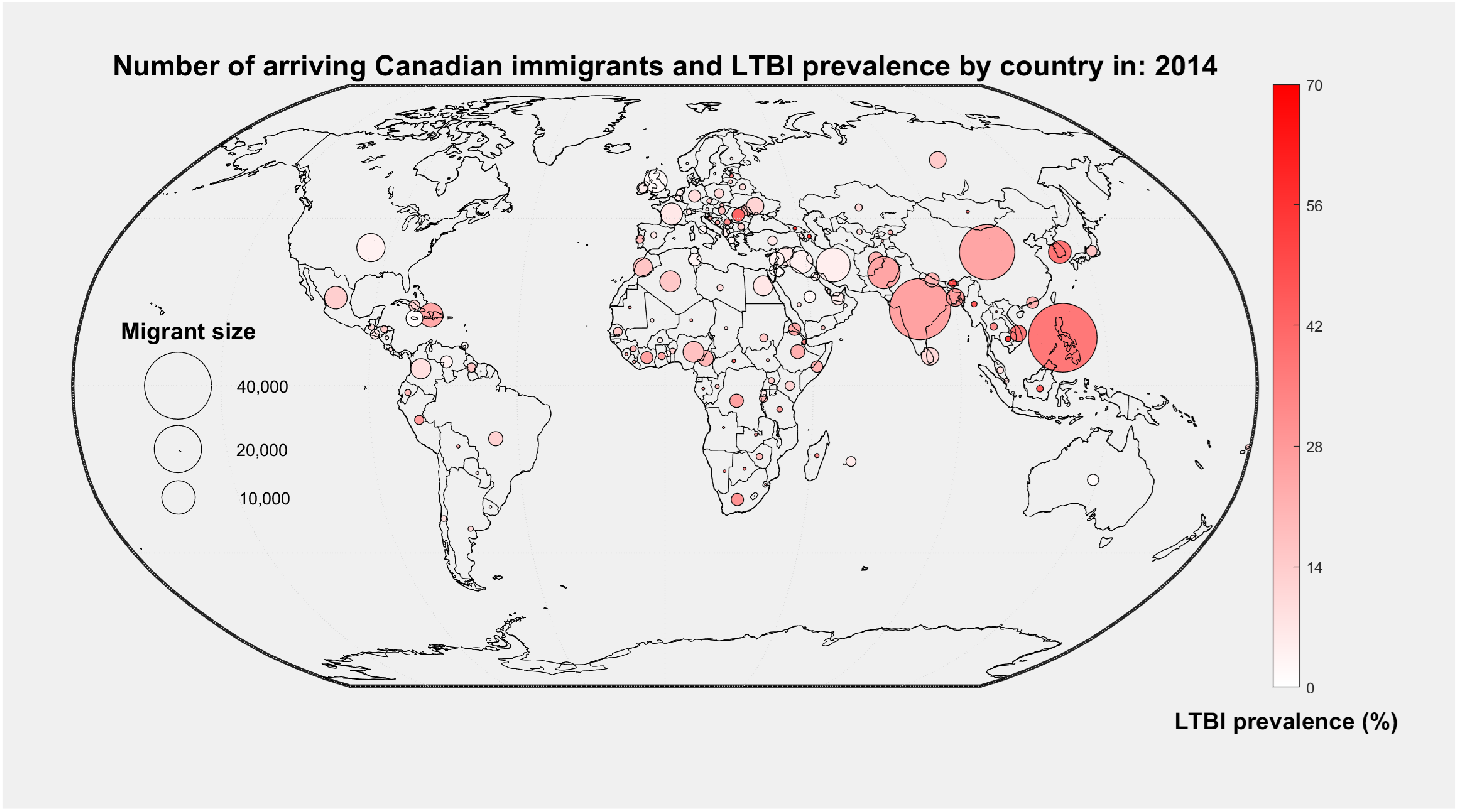
International prevalence of LTBI and immigration into Canada. Immigration numbers from various countries into Canada in 2014 visualized as area of circles. Prevalence of latent tuberculosis (estimated by Houben and Dodd (2016) (20)) corresponds to the darkness of each circle. A time series animation of estimated values between years 2000 and 2020 can be viewed at https://github.com/wruth1/Langara_Quant_Epi_Group/blob/main/Matlab%20Code/atlasmap/tb.gif.

### Immigration

Canada reports the annual number of new immigrants entering Canada, broken down by country. Both the number of new immigrants each year (21) and the number of immigrants from each of 167 countries aggregated over 5-year periods (2001-2005, …, 2016-2020) (19) is reported. Fig 2 shows immigration trends categorized by WHO geographic regions.

**Fig 2.**
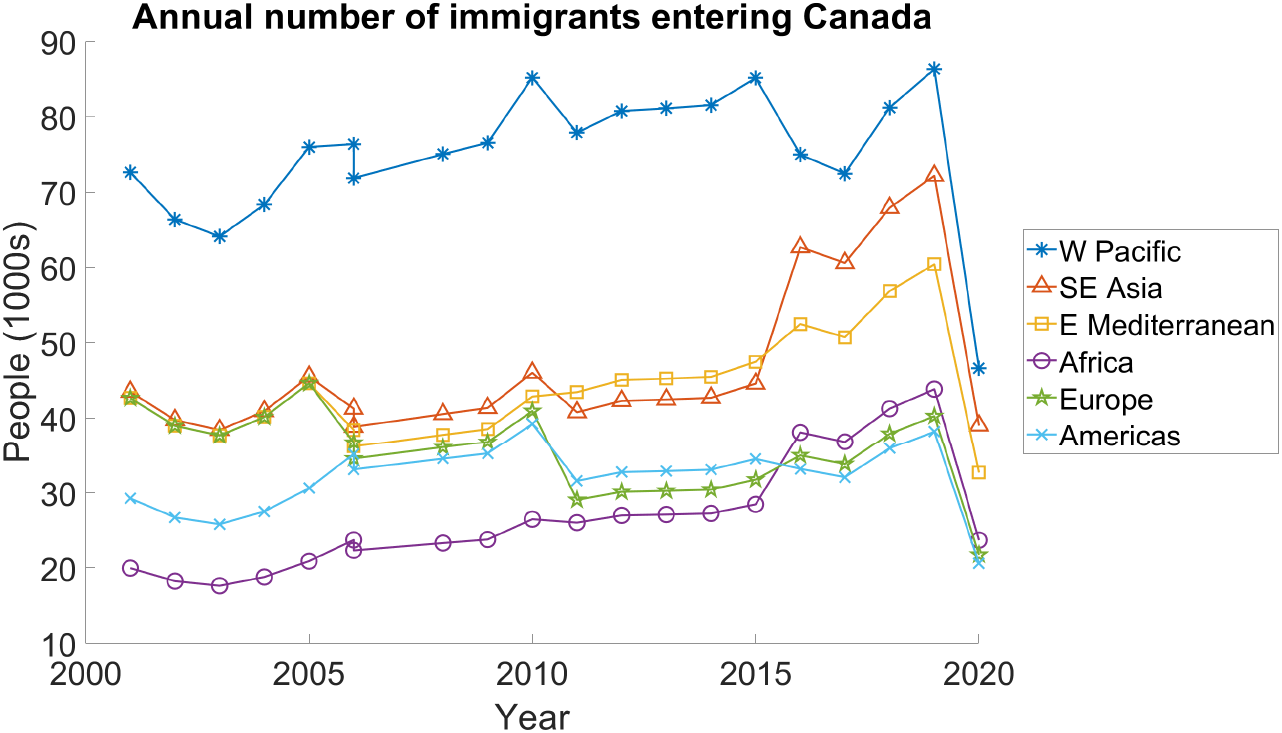
Immigration by WHO geographic region vs time. The (%) in the legend denotes the prevalence of LTBI in 2014 as reported by Houben and Dodd (2016) (20).

### Prevalence of LTBI

Houben and Dodd’s study (20) aimed to update the existing estimate of the global burden of LTBI. They found that about one-fourth of the world had LTBI, less than the previous estimate of one-third. Their study reported the prevalence of LTBI in each WHO geographic region in 2014. They also reported the prevalence of LTBI in each country in 2014, but warn that “country-level estimates are more subject to bias and uncertainty and should be used with caution”. Note that their national reports described the absolute number of LTBI cases rather than the percentage of infected individuals; we obtained a percentage by including the total population of each nation in 2014 from WHO reports (22).

### The weighted average

Recall *π*(*t*) denotes the number of new immigrants entering Canada in year *t*; for instance, *π*(2014) *≈* 260, 000. Let *π*_*c*_(*t*) denote the number of new immigrants entering Canada from country *c*. Note that *c* = 1 corresponds to Afghanistan and *c* = 167 to Zimbabwe. Assuming that each new migrant comes from only one country, it follows that for a fixed year *t*,

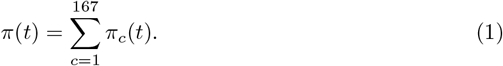

Let *q*_*LTBI*_ denote the percentage of new immigrants entering Canada with LTBI. Assuming that the number of individuals with active TB is small, and that individuals who enter Canada are uniformly randomly selected from their country of origin, then the percentage of individuals with LTBI can be estimated by

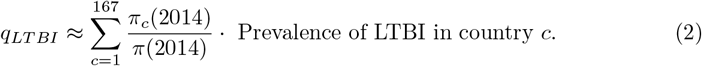

Similarly, *q*_*LTBI*_ can be estimated using Houben and Dodd’s reported prevalence by WHO geographic region (instead of by country)

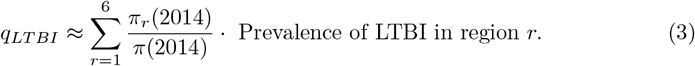

Ultimately, we estimated the burden of LTBI among new immigrants using both Houben and Dodd’s national estimates and their WHO geographic region estimates. These two estimates were nearly identical, and thus we only present estimates by WHO geographic region. Assuming that immigration data is perfect and using Houben and Dodd’s mean [95% uncertainty interval (UI)] number of individuals with LTBI (Table 1), we estimate that 22.0% [17.3%-29.0%] of immigrants entering Canada in 2014 have LTBI. Details about how we calculated this estimate is described in S1 The burden of TB among new immigrants in 2014.

**Table 1.** Prevalence of LTBI and proportion of immigrants, by WHO geographic region. The proportion of immigrants corresponds to the number of immigrants entering Canada in 2006-2021 (19), while the LTBI prevalence and recent infection mean [95% UI] are the 2014 estimate by Houben and Dodd (2016) (20).

| WHO Region | Proportion of immigrants | LTBI Prevalence (%) | Recent Infection (%) |
| --- | --- | --- | --- |
| Africa | 11.0% | 22.4 [20.6-24.6] | 1.5 [1.3-1.7] |
| Americas | 12.6% | 11.0 [7.0-20.0] | 0.2 [0.1-0.2] |
| E Mediterranean | 16.9% | 16.3 [13.4-20.5] | 0.7 [0.5-1.0] |
| Europe | 12.6% | 13.7 [9.8-19.8] | 0.3 [0.2-0.3] |
| SE Asia | 18.1% | 30.8 [28.3-34.8] | 1.2 [0.9-1.6] |
| W Pacific | 28.9% | 27.9 [19.3-40.1] | 0.5 [0.4-0.7] |

### Prevalence of recent infection

In addition to reporting the prevalence of LTBI in each WHO geographic region, Houben and Dodd (2016) report the prevalence of recent infection (20). Based on their mean [95% UI] and assuming that immigration data is perfect, we estimate that 0.71% [0.54%-0.91%] of immigrants entering Canada in 2014 had been recently infected with LTBI (within 2 years).

## Math model

Fundamentally, there are three stages of TB progression: uninfected, LTBI, and active TB infection. We present a modified SEIR (susceptible, exposed, infected, recovered) compartment model that describes how individuals move between the different stages; our model is an adaptation of the model presented by Guo and Wu (14). The model includes an immigration component that accounts for people with various stages of LTBI entering Canada. The model assumes homogeneity for individuals within each compartment.

Individuals begin their lives susceptible, represented by compartment *X*. Upon being exposed to TB, an individual moves to the “early latent” compartment, *E*. Individuals remain in *E* for an average of two years, during which they may either develop active TB, compartment *T*, or proceed to the late latent stage, *L*. The only possible progression of the infection from late latent, *L*, is to active TB, *T*. From active TB, an individual may recover, moving to compartment *R*. After recovery, the only further development our model allows is relapse into active infection, *T*, whereupon an individual may again recover, and possibly relapse again.

Each compartment also has an inflow due to immigration, although we take the rate of immigration with active TB (i.e. into compartment *T*) to be zero. We also include a constant death rate across all compartments, as well as an additional term representing the increased risk of death for individuals with active TB.

Fig 3 gives a visualization of our model, including the various compartments and transitions between them, as well as the inflows due to immigration and outflows due to death. We also group compartments based on whether they represent active TB, LTBI or non-infection.

**Fig 3.**
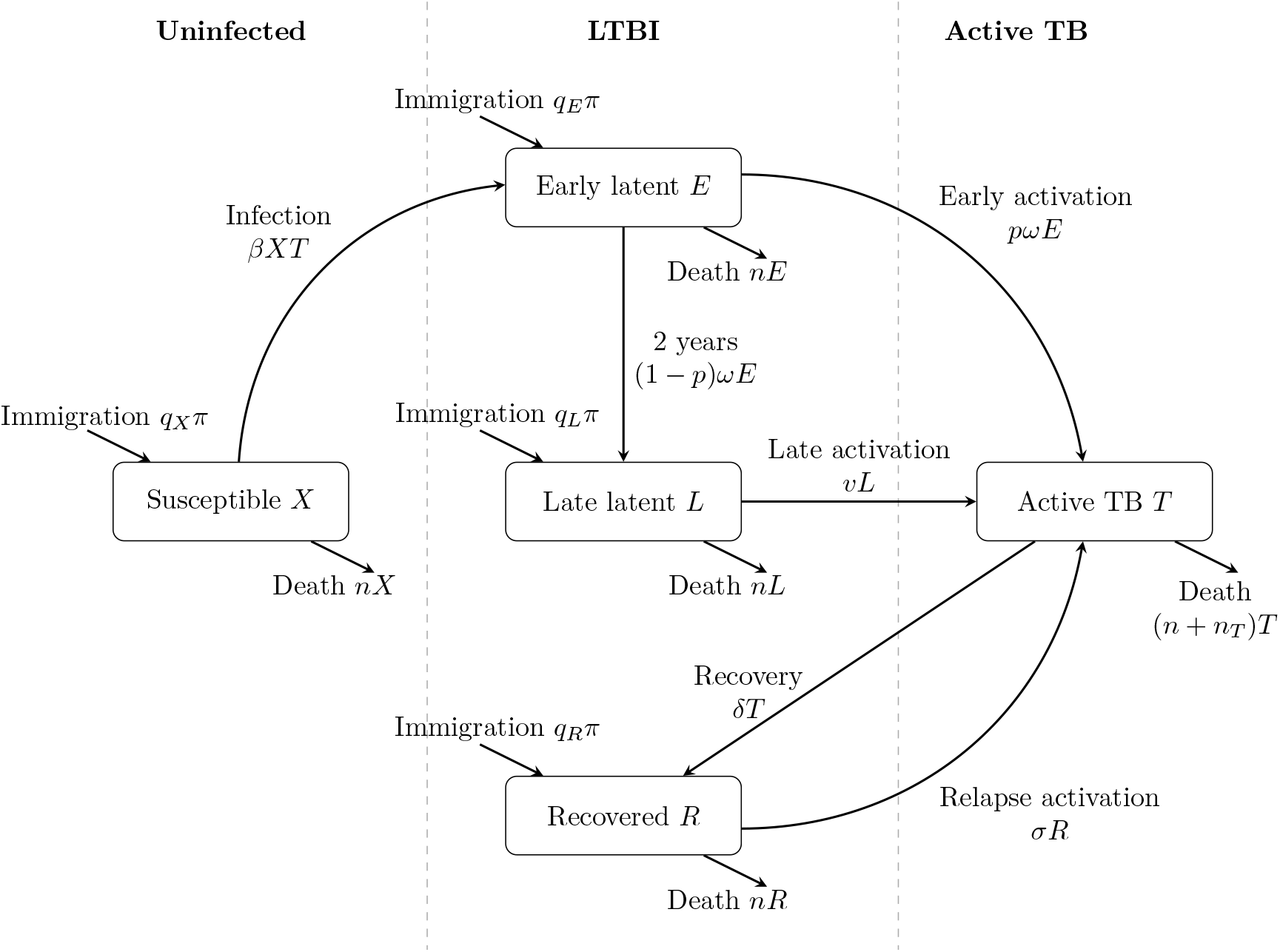
Math model of TB progression. Our model uses compartments to represent the various stages of uninfected (*X*), latently infected (*E, L*, and *R*), and actively infected (*T*) individuals. Parameters are described in Table 2.

**Fig 4.**
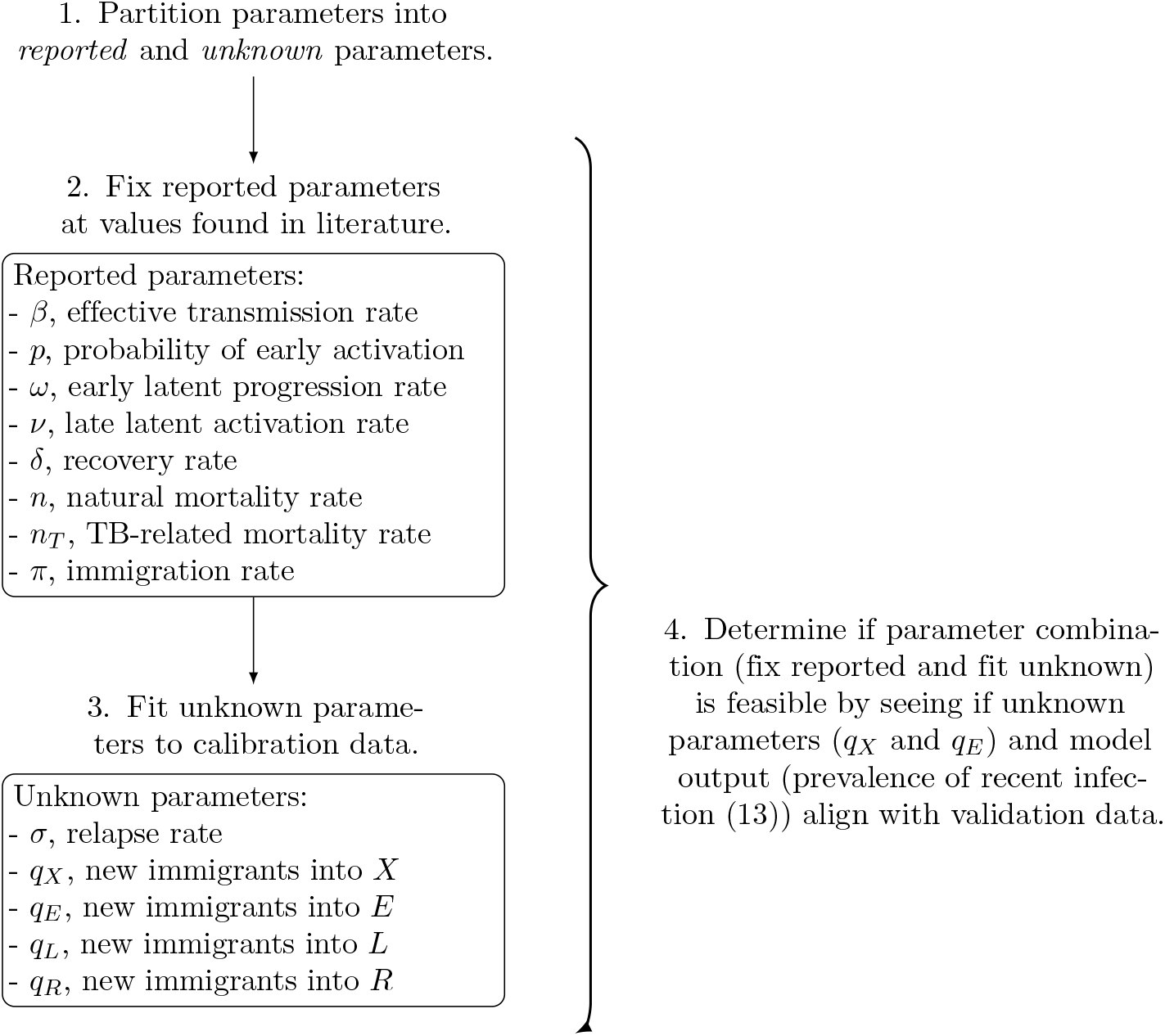
Flow diagram illustrating the algorithm used to assess the feasibility of parameter combinations in our transmission model. The calibration data and validation data are reported in Table 3.

### Transmission and latent infection

Primary transmission of the TB bacteria occurs through inhalation of the pathogen. Infectious individuals release airborne particles when they cough, talk, sneeze, or sing (23; 24).

**Table 2.** Model parameters, units, and typical values. Some parameters have large reported ranges.

| Description | Symbol | Unit | Typical value(s) | Source |
| --- | --- | --- | --- | --- |
| **Reported parameters:** |  |  |  |  |
| Effective transmission rate | $\beta$ | (person year)<sup>-1</sup> | 10<sup>-8</sup> | Guo and Wu (2011) (14) |
|  |  |  | 7 · 10<sup>-6</sup> | Ziv, Daley, and Blower (2001) (38) |
| Probability of early activation | $p$ | % | 5-15 | Colangeli et al. (2020) (39) |
|  |  |  | 5 | Menzies et al. (2018) (28) |
|  |  |  | 5, 4-10 | Jacquet et al. (2006) (30) |
| Early latent progression rate | $\omega$ | (year)<sup>-1</sup> | 0.5 | early LTBI is first 2 years (28) |
| Late latent activation rate | $\nu$ | (year)<sup>-1</sup> | 0.0017 | Menzies et al. (2018) (28) |
|  |  |  | 0.0027, 0.0011-0.0048 | Shrestha et al. (2021) (40) |
|  |  |  | 0.00040, 0.00058 | Horsburgh et al. (2010) (32) |
|  |  |  | 0.0001-0.0002 | Barnett et al. (1971) (31) |
| Recovery rate | $\delta$ | (year)<sup>-1</sup> | 0.68, 0.19-0.98 | Shrestha et al. (2021) (40) |
|  |  |  | 0.60, 0.80 | feasibility study result |
|  |  |  | 0.33 (untreated) | Tiemersma et al. (2011) (2) |
| Natural mortality rate | $n$ | (year)<sup>-1</sup> | 0.0071 | mode 2000-2021 (41) |
| TB-related mortality rate | $n_T$ | (year)<sup>-1</sup> | 0.052 | mean 2011-2020 (5) |
| Immigration rate | $\pi$ | person (year)<sup>-1</sup> | 250,000 – 350,000 | StatCan (21) |
| **Unknown parameters:** |  |  |  |  |
| Relapse rate | $\sigma$ | (year)<sup>-1</sup> | 4.6(4.2 – 5.2) · 10<sup>-4</sup> | Aldridge et al. (2016) (42) |
|  |  |  | 9.1(8.2 – 10.2) · 10<sup>-4</sup> | Aldridge et al. (2016) (42) |
|  |  |  | 0.0034 | Dowdy, Dye, and Cohen (2013) (42) |
|  |  |  | 8 · 10<sup>-5</sup> | Aparicio and Castillo-Chavez (2009) (43) |
|  |  |  | 0.05, 0.035 – 0.065 | Cohen et al. (2007) (44) |
|  |  |  | (7.49 – 7.85) · 10<sup>-5</sup> | feasibility study result |
| New immigrants into $X$ | $q_X$ | % | 78.0, 70.9 – 82.7 | synthesized estimate, Table 1 |
|  |  |  | 75.35 – 76.16 | feasibility study result |
| New immigrants into $E$ | $q_E$ | % | 0.71, 0.54 – 0.91 | synthesized estimate, Table 1 |
|  |  |  | 0.62 – 0.83 | feasibility study result |
| New immigrants into $L$ | $q_L$ | % | 6.78 – 7.35 | feasibility study result |
| New immigrants into $T$ | $q_T$ | % | 0 | assumption |
| New immigrants into $R$ | $q_R$ | % | 16.10 – 17.07 | feasibility study result |

Exposure to the bacteria may result in elimination of the pathogen either due to innate immune responses or acquired immunity (25). If an individual exposed to the bacteria fails to eliminate it, the body contains the infection in a granuloma, an aggregation of macrophages. Although the granuloma may contain the bacteria in a ‘prison’ and prevent the bacteria from spreading throughout the body and lungs, the bacteria have many cells to infect and replicate within. Individuals with granuloma containing the TB are said to have *latent tuberculosis infection*. Individuals with LTBI are not infectious and exhibit no symptoms (25). The *latent* (or *exposed)* period of an infection is defined as the time from infection to infectiousness (26; 27). Newly infected patients typically take years before developing active TB, if ever (28).

To model transmission of TB, we use the mass action transmission term described by Anderson and May (1992) (29). We define the rate of infection as

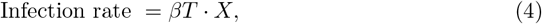

where *X* is the population of susceptible individuals, *β* the effective transmission rate, and *T* the number of actively infected individuals.

### Progression from latent to active TB infection

A granuloma may fail to contain the TB when the bacterial load becomes too great, allowing bacteria to enter the respiratory tract or bloodstream and cause infection outside the granuloma (25). When this occurs, the disease goes from being *latent* to *active*.

People who acquire LTBI are most likely to develop active TB within the first 2-years of infection (28). About 4-10% of otherwise healthy adults will develop active TB within 2 years of being exposed (28; 30). After 2 years, people with LTBI are far less likely to develop active TB. For those with stable LTBI, the annual probability of developing active TB is reported variously as 0.01-0.02% (31), 0.040-0.058%(32), or 0.17% (28).

To model fast vs slow progression, we define two additional compartments: the early latent compartment *E* (individuals who have been latently infected for less than 2 years) and the late latent compartment *L* (individuals who have been latently infected for more than 2 years and have never had active TB). We define *fast-progression* as an individual proceeding directly from the early latent stage to active TB, and *slow-progression* as an individual proceeding from early latent through late latent to active TB.

By definition, the combined out-flow from *E* (aside from death) is given by

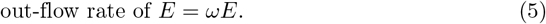

Our model assumes each compartment is homogeneous, and cannot distinguish when individuals entered the compartment; as such, we assume that *ω* = 0.5 so that the average time that individuals spend in *E* is 2 years.

People in *E* who do not die will either progress to late latent *L* or develop active TB and enter *T*. If *p* is the probability a latently infected individual develops active TB within 2 years of infection, then

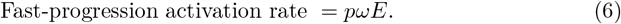

It follows that individuals in *E* enter *L* at rate (1 *− p*)*ωE*.

Similarly, we define

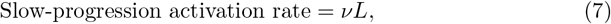

Slow-progression activation rate = ν*L*, (7) where ν is the annual probability someone in *L* develops active TB.

### Active TB, recovery, and death

We assume all compartments have the same natural mortality rate *n*. The active TB compartment *T* has an additional death term *n*_*T*_ that describes death due to active TB.

People who do not die recover at rate

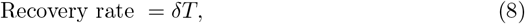

where δ is the recovery rate. Note that the reciprocal of δ gives the average duration an individual has active *T B* before recovering.

#### Developing active TB more than once

People who have recovered from active TB are not immune from developing active TB again (6; 33; 34). Individuals who develop active tuberculosis more than once in their lifetime can be classified into either *endogenous relapse* or *exogenous reinfection* (33). Two tuberculosis treatment trials conducted in the United States and Canada found that 96% of recurrent tuberculosis cases were from reactivation of the initial infecting strain (i.e., relapse) (35). As such, we assume that there is no reinfection taking place in Canada.

We define the

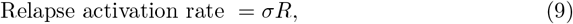

where *σ* is the annual probability of relapse active TB.

### Immigration

Statistics Canada (StatCan) reports the annual number of new immigrants entering Canada (21). We define the immigration term *π*(*t*) to be the number of new immigrants entering Canada in year *t*.

To model how immigration affects the burden of TB, we additionally define *q*_*X*_ to be the percentage of new immigrants that enter Canada into the *X* compartment. We similarly define *q*_*E*_, *q*_*L*_, *q*_*T*_, and *q*_*R*_ to respectively denote immigrants that enter *E, L, T*, and *R*.

*Screening for active TB*. All permanent migrants entering Canada are required to undergo chest radiography to be screened for active tuberculosis disease (36). Many additional groups are required to undergo other forms of active TB screening before being admitted to Canada (e.g. individuals who are on extended stays of over 6 months, come from high incidence countries, or have healthcare-related occupations). Thus, we assume that no immigrants entering Canada have active TB; i.e.,

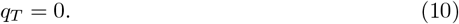

In Canada, LTBI screening is not performed en masse for migrant groups pre-entry, and is instead performed on select migrants who are at higher risk to having been exposed to TB infection (36; 37). The incidence rate in the country of origin, proximity to active cases, and occupation are factors used to assess risk and selection for LTBI screening. Those who test positive for LTBI are still allowed entry, but are subject to medical surveillance (36). LTBI tests are still discouraged for mass screening migrants due to its cost:benefit ratio, and reserved for high risk travellers (37). As such, we are not aware of any reliable sources for the values of *q*_*X*_, *q*_*E*_, *q*_*L*_, and *q*_*R*_.

### Equations

Taken together, the assumptions discussed above give rise to the following system of differential equations.

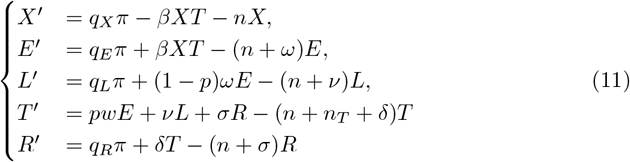

Fig 3 gives a diagram of our compartments, along with arrows and rates for all possible transitions. Table 2 gives more detail on each of the parameters in our model.

Solving this system of differential equations gives the number of susceptibles *X*(*t*) at time *t*, early latently infected *E*(*t*), late latently infected *L*(*t*), active TB *T* (*t*), and recovered *R*(*t*). With these, we can compute key epidemiological quantities like the incidence of active TB *pwE* + *vL* + *σR*, prevalence of active TB *T*, and prevalence of LTBI *E* + *L* + *R*. More generally, our model allows us to compute other quantities that can be compared to reported values in the literature.

### Model calibration and estimates

Our model contains many parameters that describe the epidemiology of tuberculosis in Canada. Genomic studies demonstrate that globally, *M. tuberculosis* has considerable genetic variability, which lead to varying levels of infectivity, drug resistance, and deadliness (25). Hence, reported parameter values may have large ranges. Table 2 summarizes all of our model’s parameters, including their units and typical values.

### Finding feasible parameters

Parameters are broadly divided into two groups: those obtained from the literature (the *reported* parameters), and those we estimate (the *unknown* parameters). We begin by fixing all reported parameters at plausible values. We then estimate the unknown parameters by fitting to our calibration data (this fitting process is more precisely described in S3 Fitting unknown parameters to calibration data).

To determine whether a parameter combination (i.e., the fixed reported parameters and estimated unknown parameters) is feasible, our model computes other quantities that are compared to validation data. If the quantity computed from our model is incompatible with the validation data (defined by being within the *plausible range*, which correspond to reported data in Table 3), we discard the parameter combination and call it *infeasible*. If all model computed quantities are within the plausible ranges, we say the parameter combination is *feasible*. Table 3 summarizes the calibration data, the validation data (and the plausible range to be “close” to the validation data), and additional data we compare our model to.

**Table 3.**
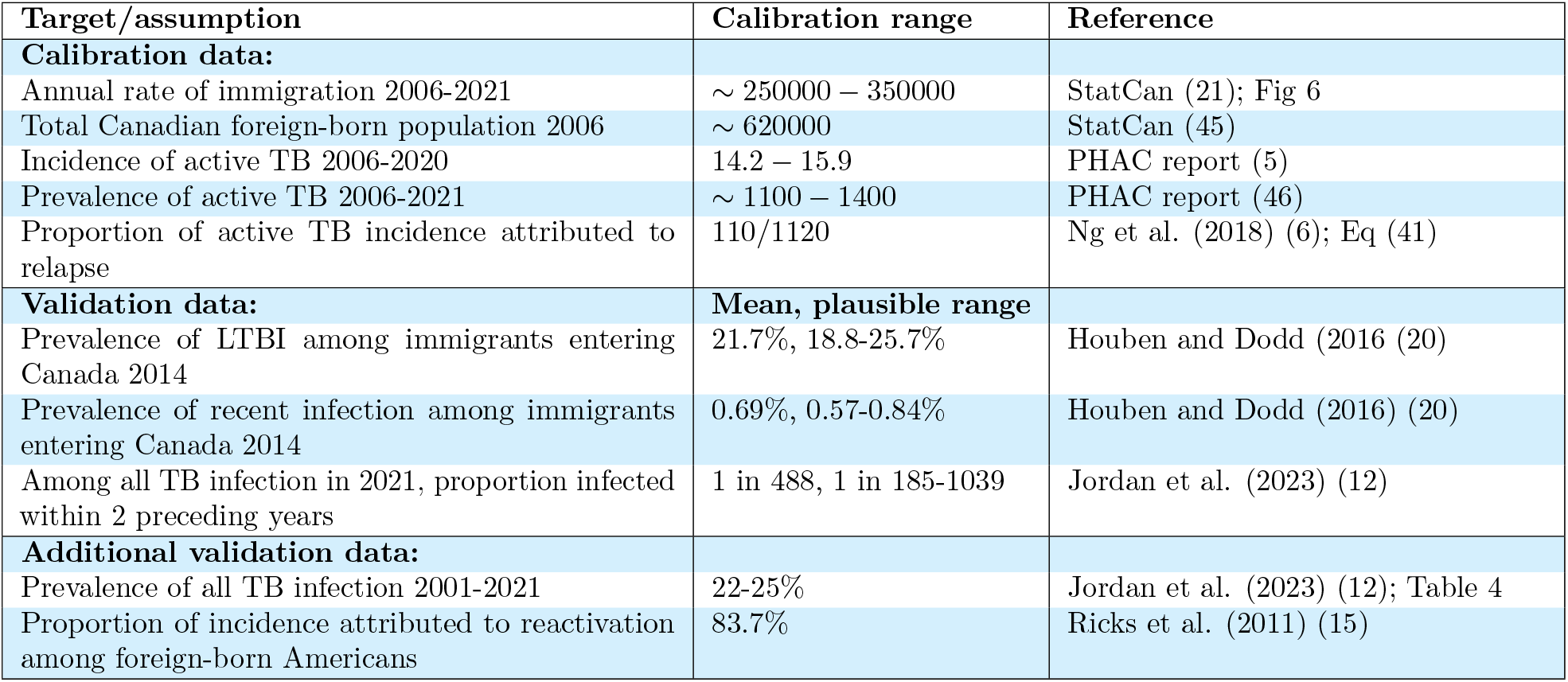
Data used by our model. Calibration targets and demographic assumptions about Canadian foreign-born population. Unless otherwise stated, the incidence (per 100,000 individuals) and prevalence is among foreign-born Canadians.

### Validation data

*Prevalence of LTBI among new immigrants*. We can compare our model’s output to our estimate of *q*_*LT BI*_, the percentage of new immigrants with LTBI (this weighted average is described in Estimating the burden of TB among new Canadian immigrants). Recall that *q*_*X*_ (*q*_*T*_) is the percentage of new immigrants that are uninfected (actively infected). Due to screening, we assumed that *q*_*T*_ = 0, and since *q*_*X*_ + *q*_*E*_ + *q*_*L*_ + *q*_*T*_ + *q*_*R*_ = 100%, it follows that

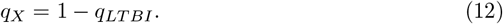

Recall *q*_*LT BI*_ is estimated by Eq (2) or (3). The quantity *q*_*X*_ can be computed by our model; if *q*_*X*_ is outside the plausible range, we consider this solution infeasible.

*Burden of TB among foreign-born Canadians*. The Public Health Agency of Canada (PHAC) regularly releases reports that summarize the descriptive epidemiology of active TB in Canada (5; 46), including the annual incidence of active TB among foreign-born Canadians in each year, and an estimate of the prevalence of active TB. This data is summarized in Fig 5; we compare our model results to this data in Fig 7.

**Fig 5.**
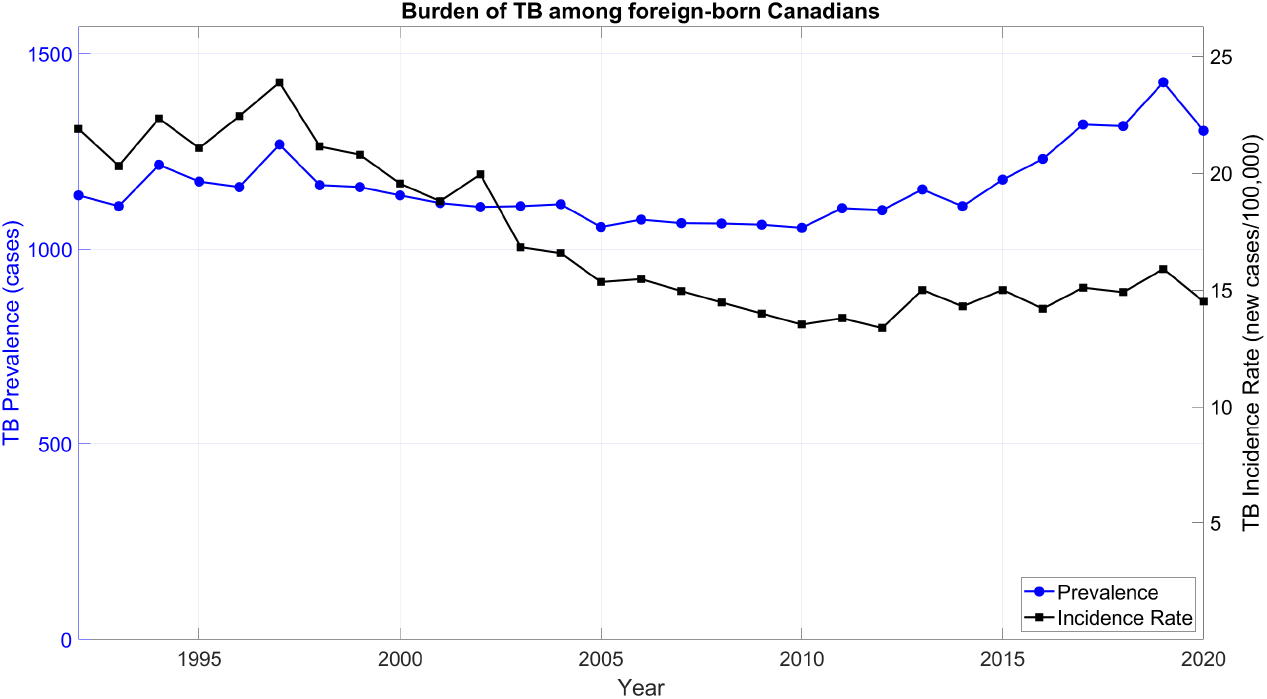
Reported incidence and prevalence. The burden of TB among foreign-born Canadians from 1992 to 2020, as disclosed by the PHAC in their 2002, 2012, and 2022 reports (5; 46; 47).

**Fig 6.**
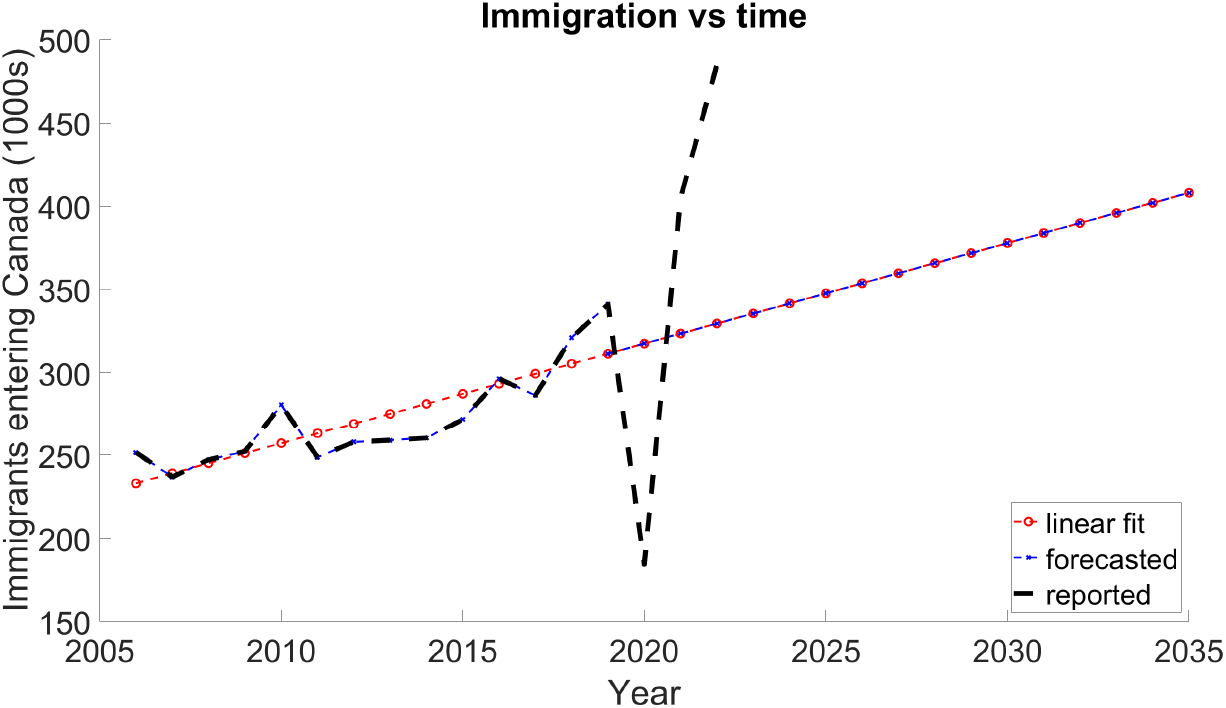
Immigration vs time. Reported and forecasted immigration levels.

**Fig 7.**
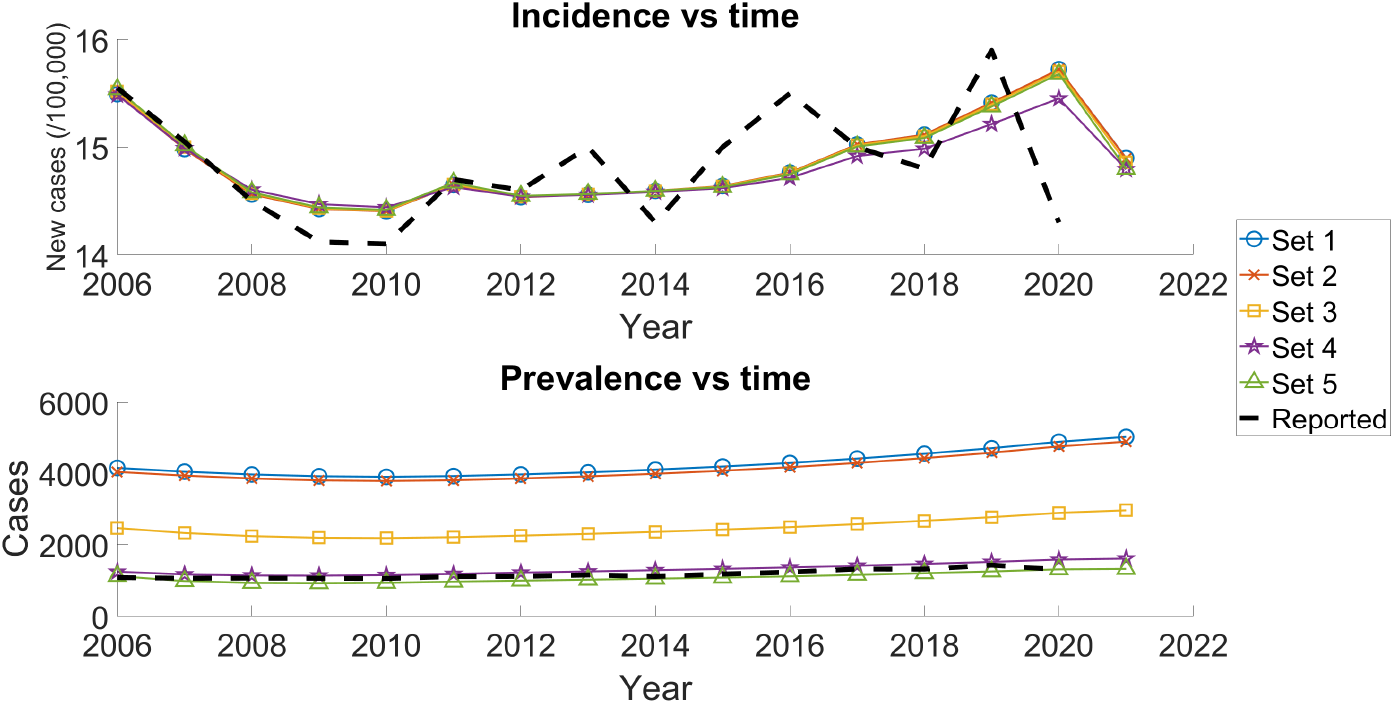
Model estimates of incidence and prevalence. Each trajectory corresponds to one of the 5 feasible parameter sets (Table 5).

*Prevalence of LTBI*. Our model includes the prevalence of LTBI among new immigrants, as shown in Fig 9. In Canada, the case count of LTBI is not reported by the PHAC (5). Instead, we compare our estimates to another author’s reported findings. Jordan et al. (2023) estimated the prevalence of TB infection among the Canadian foreign-born population in 2001, 2006, 2011, 2016, and 2021 (see Table 4). They also report that among all foreign-born Canadian residents with tuberculosis infection in 2021, only 1*/*488 *≈* 0.2% (95% UI 1/1039 to 1/185) had become infected within the preceding 2 years, which can be estimated as

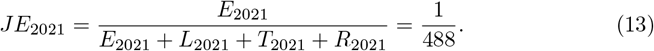

**Table 4.** Prevalence of TB infection among foreign-born Canadians. Jordan et al. (2023) estimates of the prevalence of TB among foreign-born residents in Canada (12).

| Year | Latent or active TB prevalence | 95% confidence interval |
| --- | --- | --- |
| 2001 | 25% | 20-35% |
| 2006 | 24% | 20-33% |
| 2011 | 23% | 19-30% |
| 2016 | 22% | 19-28% |
| 2021 | 22% | 19-27% |

**Table 5.** Combinations of parameters that satisfied calibration ranges. The unknown parameters (*σ, q*_*E*_, *q*_*L*_, *q*_*R*_, *X*_0_, *E*_0_, *L*_0_, *T*_0_, and *R*_0_) are the fit parameters; all other parameters were fixed at a reported value. The only reported parameters that had multiple feasible values were the TB-induced death rate *n*_*T*_ and active TB recovery rate δ.

| Parameter | Set 1 | Set 2 | Set 3 | Set 4 | Set 5 |
| --- | --- | --- | --- | --- | --- |
| $\beta$ | $1.0 \times 10^{-8}$ | $1.0 \times 10^{-8}$ | $1.0 \times 10^{-8}$ | $1.0 \times 10^{-8}$ | $1.0 \times 10^{-8}$ |
| $p$ | $2.0 \times 10^{-1}$ | $2.0 \times 10^{-1}$ | $2.0 \times 10^{-1}$ | $2.0 \times 10^{-1}$ | $2.0 \times 10^{-1}$ |
| $\omega$ | $5.0 \times 10^{-1}$ | $5.0 \times 10^{-1}$ | $5.0 \times 10^{-1}$ | $5.0 \times 10^{-1}$ | $5.0 \times 10^{-1}$ |
| $\nu$ | $1.7 \times 10^{-3}$ | $1.7 \times 10^{-3}$ | $1.7 \times 10^{-3}$ | $1.7 \times 10^{-3}$ | $1.7 \times 10^{-3}$ |
| $n_T$ | $5.2 \times 10^{-2}$ | $6.0 \times 10^{-2}$ | $5.2 \times 10^{-2}$ | $5.2 \times 10^{-2}$ | $5.2 \times 10^{-2}$ |
| $\delta$ | $2.0 \times 10^{-1}$ | $2.0 \times 10^{-1}$ | $4.0 \times 10^{-1}$ | $8.0 \times 10^{-1}$ | $1.0 \times 10^0$ |
| $n$ | $7.1 \times 10^{-3}$ | $7.1 \times 10^{-3}$ | $7.1 \times 10^{-3}$ | $7.1 \times 10^{-3}$ | $7.1 \times 10^{-3}$ |
| $\sigma$ | $7.50 \times 10^{-5}$ | $7.49 \times 10^{-5}$ | $7.85 \times 10^{-5}$ | $7.72 \times 10^{-5}$ | $7.77 \times 10^{-5}$ |
| $q_X$ | 75.36% | 75.35% | 76.16% | 75.87% | 76.05% |
| $q_E$ | 0.80% | 0.80% | 0.81% | 0.62% | 0.83% |
| $q_L$ | 6.78% | 6.78% | 6.93% | 7.35% | 6.97% |
| $q_R$ | 17.06% | 17.07% | 16.10% | 16.16% | 16.15% |
| $X_0$ | 4,720,005 | 4,719,658 | 4,770,023 | 4,693,091 | 4,755,816 |
| $E_0$ | 5,163 | 5,160 | 5,068 | 4,016 | 5,056 |
| $L_0$ | 204,860 | 205,023 | 211,333 | 272,138 | 212,667 |
| $T_0$ | 4,138 | 4,029 | 2,462 | 1,243 | 1,122 |
| $R_0$ | 1,252,783 | 1,253,080 | 1,198,063 | 1,216,462 | 1,212,290 |

**Table 6.** Forecasted *End TB Strategy* 2035 incidence goals. To meet these goals, Canada should have a 90% reduction in TB incidence compared with 2015 (15.0 per 100,000). Our model shows that we will not meet these goals under various assumptions. The values for LTBI prevalence are those reported by Houben and Dodd (20) (Table 1).

| Immigration | LTBI prevalence | Incidence in 2035 | Compared to 2015 |
| --- | --- | --- | --- |
| *Goal* | - | *1.50* | *-90.0%* |
| Baseline | high | 18.3 | +22.0% |
|  | mid | 14.6 | -2.7% |
|  | low | 12.2 | -18.7% |
| None | low | 8.3 | -44.7% |

Fig 10 compares our model estimate of the prevalence of LTBI to those of Jordan et al.

#### Proportion of incidence attributed to reactivation

Ricks et al. (2011) studied 22,151 cases of active TB from foreign-born individuals, genotyped by the U.S. National TB Genotyping Service in 2005-2009 (15), of which 18,540 (83.7%) were reactivations. We consider progression from both late latent *L* and recovered *R* into active *T* to be a reactivation, so

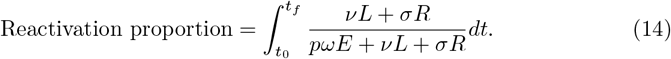

### Estimating the active TB case detection rate

Notice that

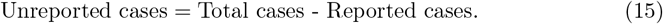

Our model can estimate total cases (i.e., the prevalence of active TB); comparing this to the reported case count (5) allows us to estimate the number of unreported cases. Thus, we estimate the case detection rate with the *underreport proportion* defined as

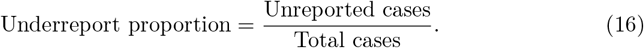

As demonstrated in Fig 7 and Table 7, our model’s estimate of total prevalence is very sensitive to the recovery rate. Note that the reciprocal of the recovery rate is the average duration of active TB. Fig 8, we demonstrate the underreport percentage as a function of the average duration of active TB. Note that negative numbers may occur because in Eq (15), the reported cases could be larger than the model estimate.

**Table 7.**
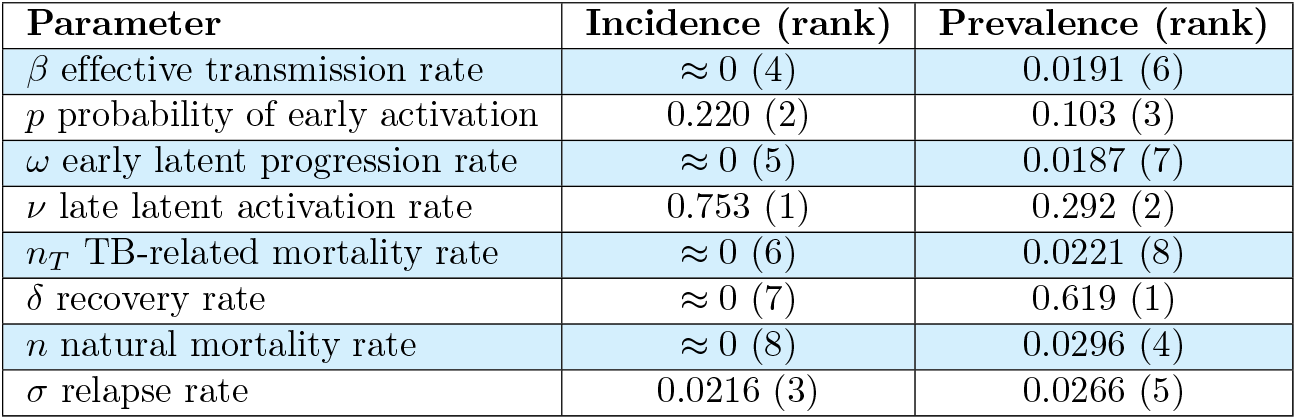
Sensitivity analysis results. We determine the sensitivity of active TB incidence (and prevalence) in 2035 to our parameters by computing the total-effect indices (49). These estimates correspond to using *~* 1, 000, 000 Monte Carlo samples.

| Parameter | Incidence (rank) | Prevalence (rank) |
| --- | --- | --- |
| $\beta$ effective transmission rate | $\approx 0$ (4) | 0.0191 (6) |
| $p$ probability of early activation | 0.220 (2) | 0.103 (3) |
| $\omega$ early latent progression rate | $\approx 0$ (5) | 0.0187 (7) |
| $\nu$ late latent activation rate | 0.753 (1) | 0.292 (2) |
| $n_T$ TB-related mortality rate | $\approx 0$ (6) | 0.0221 (8) |
| $\delta$ recovery rate | $\approx 0$ (7) | 0.619 (1) |
| $n$ natural mortality rate | $\approx 0$ (8) | 0.0296 (4) |
| $\sigma$ relapse rate | 0.0216 (3) | 0.0266 (5) |

**Table 8.** The reported parameters are fixed at values similar to those reported in the literature. The values explored for the unknown parameters are the initial conditions used for the optimizer.

| Parameter | Values explored |
| --- | --- |
| **Reported:** |  |
| $\beta$ | $1.0 \times 10^{-8}, 7.0 \times 10^{-6}$ |
| $p$ | 0.08, 0.10, 0.15, 0.20, 0.0154 |
| $\omega$ | 0.50 |
| $v$ | $1.0 \times 10^{-4}, 2.0 \times 10^{-4}, 5.0 \times 10^{-4}, 1.7 \times 10^{-3}$ |
| $n_T$ | 0.0522, 0.06 |
| $\delta$ | 0.20, 0.40, 0.60, 0.80, 1.00 |
| $n$ | 0.0071 |
| **Unknown:** |  |
| $\sigma$ | $8.0 \times 10^{-5}, 3.0 \times 10^{-3}$ |
| $q_E$ | 0.563%, 0.92% |
| $q_L$ | 10% |
| $q_R$ | 10% |
| $X_0$ | 2, 435, 279 |
| $E_0$ | 14, 846 |
| $L_0$ | 3, 096, 493 |
| $T_0$ | 1081 |
| $R_0$ | 639, 250 |

**Fig 8.**
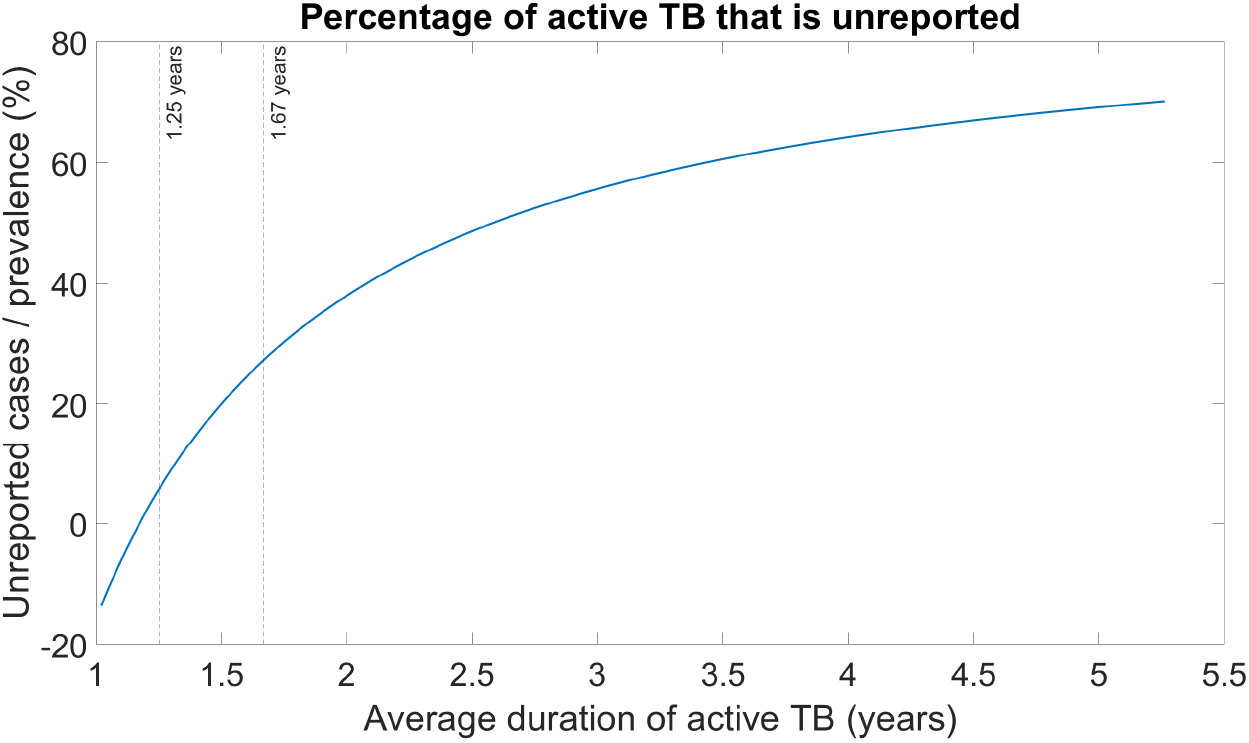
Unreported cases vs average infectious period. The range of average duration of active TB comes from values reported by (40). The lines at infectious periods of 1.67 and 1.25 correspond to the average duration of active TB found in our feasibility study; they respectively had an underreport percentage of 27% and 6%, and thus a case detection rate of 73% and 94%. Negative underreport percentages would imply overreporting.

**Fig 9.**
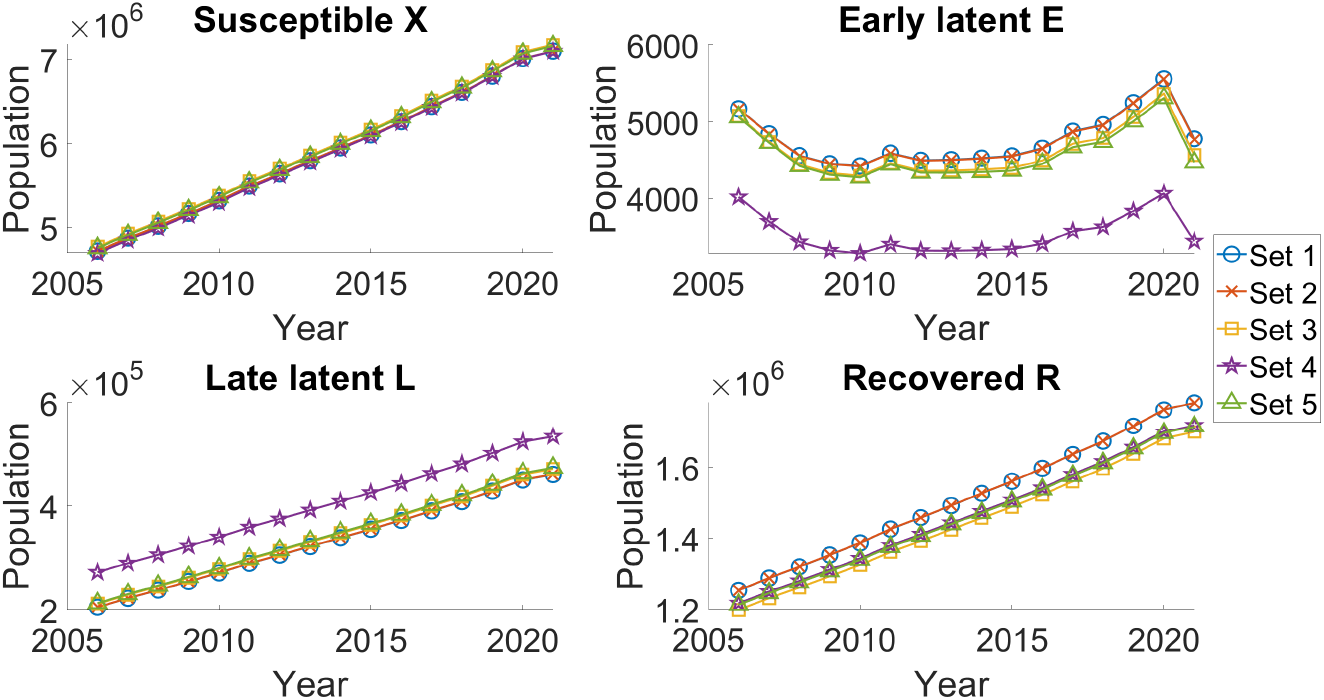
Trajectories of compartment sizes vs time. Each trajectory corresponds to one of the 5 feasible parameters (Table 5). Reduced immigration in 2020 drives a drop in *E*.

**Fig 10.**
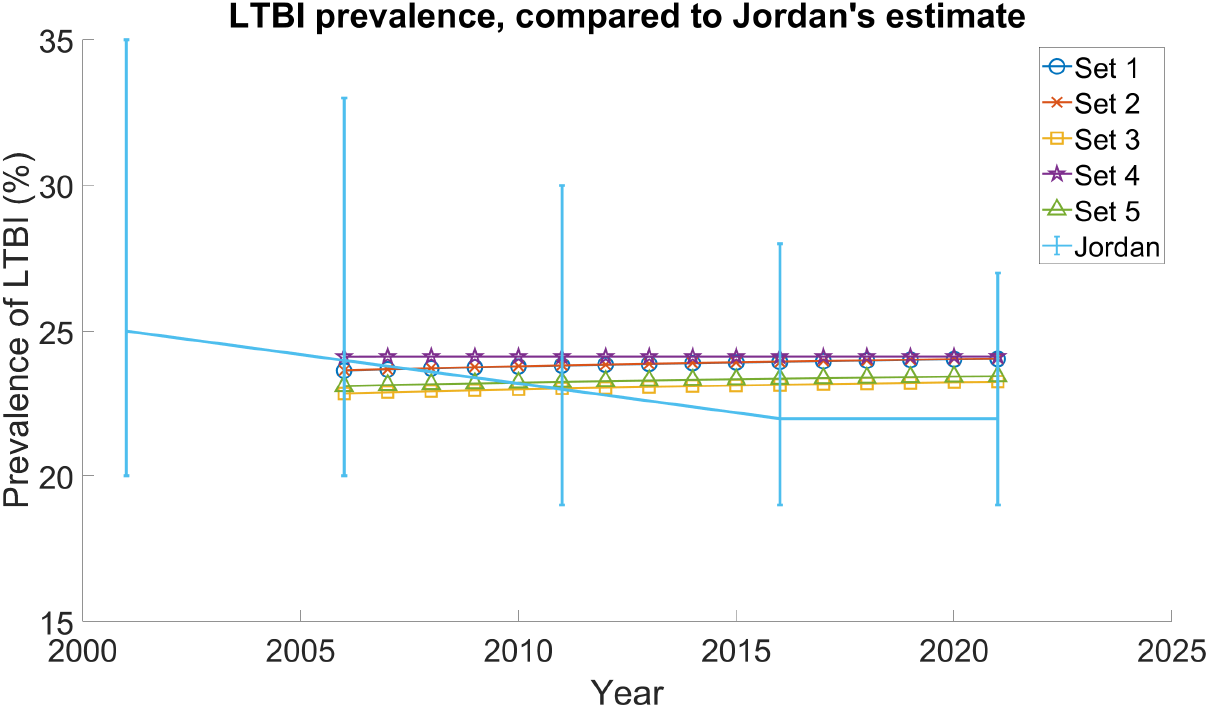
Prevalence of LTBI among foreign-born Canadians. Each trajectory is based on each of the four feasible parameter sets (see Table 5). For comparison, we also display the estimate (and 95% UI) by Jordan et al. (2023); our estimate was *not* trained on Jordan’s data, but we obtained very similar results.

### Estimating the burden of TB in 2035

To predict the incidence of active TB in 2035, we need to forecast the future total number of new immigrants, which WHO geographic regions or countries new immigrants arrive from, and the burden of TB among new immigrants.

#### Extrapolating immigration

We extrapolate the annual rate of immigration using a linear model fit to historical data. Due to the COVID-19 pandemic and subsequent government responses, we omit data from 2020 through 2023. We fit our predictor using least-squares regression. See Fig 6 for a plot of our regression model alongside actual data. We see that the regression fits historical data quite well.

Denote the reported annual immigration by *π*_*R*_(*t*) and the prediction made by our linear regression by *π*_*L*_(*t*). To make full use of historical data, we define the extrapolated immigration function

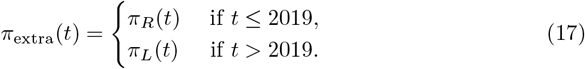

In addition to the total number number of new immigrants, we need to specify which WHO geographic regions (or countries) immigrants are coming from. We investigate four scenarios: *baseline*, where the proportion of immigrants from each WHO geographic region matches the average from 2006-2021 (see Table 1); *worst incidence* or *SEA*, where we assume immigration occurs only from the highest incidence WHO geographic region (SEA); *best incidence* or *AMR*, where we assume immigration occurs only from the lowest incidence WHO geographic region (AMR); and *no immigration* or *none*, where we assume there is no additional immigration after 2021.

#### Burden of TB among new immigrants

Our model requires the burden of TB infection among new immigrants; i.e., parameters *q*_*X*_, *q*_*E*_, *q*_*L*_, *q*_*T*_, and *q*_*R*_. We again assume that Canada again continues to screen for active TB, and thus *q*_*T*_ = 0. In order to to investigate the impacts of immigration from different WHO geographic regions, we no longer assume these parameters are constant in time. In S5 Forecasting the burden of LTBI among new immigrants, we describe the estimation of *q*_*X*_, *q*_*E*_, *q*_*L*_, and *q*_*R*_ based on the assumption of their WHO geographic region of origin.

*Global prevalence of LTBI*. To forecast the burden of TB among new immigrants, our model requires forecasting the global prevalence of LTBI. In addition to their estimates in 2014, Houben and Dodd (2016) applied their methodology in 1997 and estimated the global burden of LTBI then was 26.9% [95% UI 22.4%-32.7%] (20).

Let *P*(*t*) (%) denote the global prevalence of LTBI. In recent years, it has been reported that the annual TB incidence has been decreasing at an exponential rate (25; 48), and so we assume that the global prevalence of LTBI is also decaying at an exponential rate. This gives

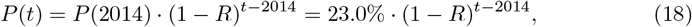

where *R* is the annual rate of prevalence decay. Evaluating *t* = 1997 and solving for *R* gives

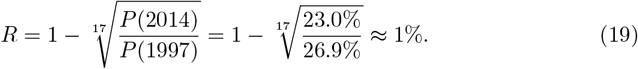

This prevalence estimate is reasonably in line with other estimates of the global incidence of TB: Pai et al. (2016) estimate the annual TB incidence is decreasing each year by 1.5% (25), while reports from 2010-2020 show the annual rate of TB incidence dropped an average of 2.4% a year (48).

Our model requires the prevalence of TB in each WHO geographic region. Following Eq (19), we assume the prevalence of each region decays at 1% per year. So for instance, using data from Table 1, the prevalence of LTBI in AFR at year *t* would be

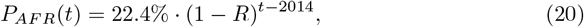

where *R* = 1%.

#### Sensitivity analysis

To study the sensitivity of model output to changes in parameter values, we used global sensitivity analysis methods based on the work of Saltelli (2002) and Saltelli et al. (2008) (49; 50). Specifically, we use the total-effect index. Briefly, this index measures the sensitivity of a model output to changes in one of its inputs. Values range from zero to one, with values close to one corresponding to stronger association, and values close to zero indicating weaker association.

We chose two outputs to examine: the incidence and prevalence of active TB in 2035. We compute the sensitivity of both outputs to each of the reported parameters (see Table 2). The results of the sensitivity analysis are displayed in Table 7. Details about how we compute them is described in S4 Sensitivity analysis.

## Results and discussion

### Feasible parameters

Following the feasibility study (summarized in Table 3), we found four feasible parameter combinations; these are presented in Table 5. Notice that the relapse activation rate, *σ* (which was not a parameter compared to any validation data) is comparable to other values in the literature (Table 2).

We found that the only feasible value of the early activation probability was *p* = 20%, which is larger than its typical values (see Table 2). We suspect this may be because immigration to a new country is generally quite stressful, and stress has been linked to the activation of latent TB (51; 52).

### Incidence and prevalence of active TB

Fig 7 demonstrates how our model’s estimates of incidence and prevalence of active TB compares to their reported values. Note that our model was calibrated directly to the incidence and indirectly to the prevalence (this fitting procedure is explained in detail in S3 Fitting unknown parameters to calibration data). Thus, it is unsurprising that our model does very well at predicting the incidence.

Our model generally overestimates the prevalence. This is reasonable since reports on active TB are often underreported – the WHO estimates that over one-third of all individuals who develop active TB diseases are never diagnosed nor counted by public health authorities (48). Note that the Canadian report does not make any effort to adjust for underreporting, and only includes confirmed cases of active TB (laboratory confirmed or clinically diagnosed) (5).

Fig 8 shows our model’s estimate of the case detection rate of active TB among the foreign-born Canadian vs the average duration of active TB. Our model estimated the recovery rate of active TB at 0.6 and 0.8 (respectively corresponding to infectious periods of 1.67 and 1.25 years), resulting respectively in case detection rates of 73% and 94%. These values are higher than WHO’s global estimate of 67% (48), which is reasonable given Canada’s significant investment in TB monitoring (3; 4; 5; 6).

### Prevalence of LTBI

Using the feasible parameters (Table 5), we compute the prevalence of LTBI. As demonstrated in Fig 10, our results are consistent with estimates from Jordan et al. (2023) (12).

*Proportion of incidence attributed to reactivation*. Again using our feasible parameter combinations, we compute Eq (14) and found that the reactivation proportion among foreign-born Canadians was 58.1% and 69.3%, which is comparable to the reported 83.7% among foreign-born Americans (15).

### The burden of TB in 2035

WHO’s *The End TB Strategy* aims for a 90% reduction by 2035 in TB incidence rate compared with 2015 (53). In 2015, the incidence of active TB among foreign-born Canadians was 15.0 per 100,000 (5), and thus the goal is an incidence of 1.50 per 100,000 in 2035.

Assuming the *baseline* scenario (immigrants arrive in the same proportions from each WHO geographic region as 2006-2021) and assuming the global prevalence of LTBI is within the 95% UI of Houben and Dodd (20) (presented in Table 1), our model predicts that among foreign-born individuals, Canada will substantially fail to meet the WHO’s target of 90% reduction by 2035.

To estimate the incidence of active TB attributed to latent activation of the current foreign-born population of Canada, we examine our model assuming that no new immigrants enter Canada from now onward. In this scenario, assuming the lowest global prevalence of LTBI, Canada will still fail to reach the 90% reduction. We therefore conclude the reservoir of latently infected individuals activating will cause Canada to fail to reach its incidence goals. Trying to reach the 90% reduction goal by controlling future immigration is not a plausible solution – instead, we need to address the pool of latently infected individuals, perhaps by testing and treating LTBI.

Fig 11 demonstrates the forecasted trajectories of incidence of active TB under a variate of assumptions, while Table 6 summarizes the projected incidence in 2035 compared to the incidence in 2015.

**Fig 11.**
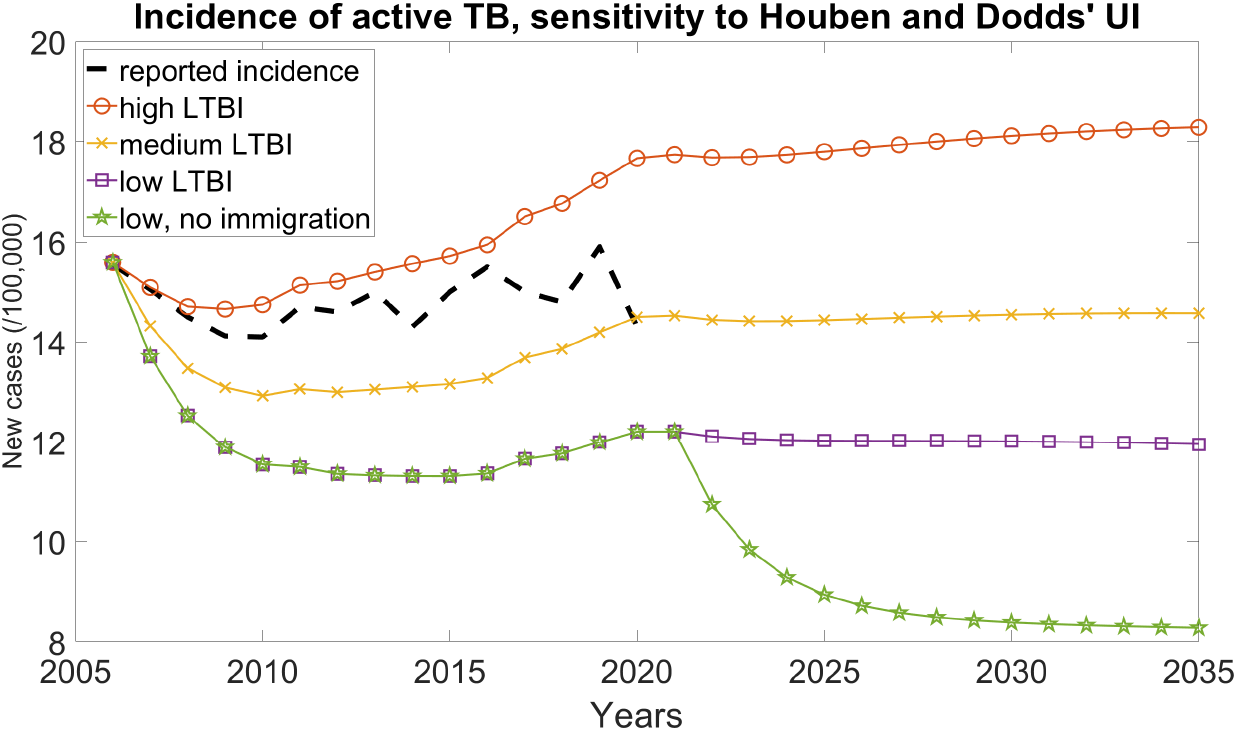
Forecast of TB incidence in 2035. WHO’s *The end TB strategy* aims for a 90% reduction by 2035 in TB incidence rate compared with 2015 (53). If current immigration trends continue, regardless of which WHO geographic region Canada allows immigrants to enter from, our model predicts that Canada will substantially fail to reach the 2035 incidence goal of 1.50 per 100,000.

#### Forecasted prevalence

Recall we use our model to simulate the dynamics of TB up to 2035 assuming various scenarios about future immigration, including immigration demographics matching those from 2006-2020, all immigration from the highest-incidence WHO geographic region (SEA), and all immigration from the lowest-incidence WHO region (AMR). Under these scenarios, the forecasted prevalence of active and latent TB in 2035 are respectively displayed in Fig 12 and Fig 13. As expected, admitting immigrants from regions with a high TB burden increases the prevalence of TB in Canada.

**Fig 12.**
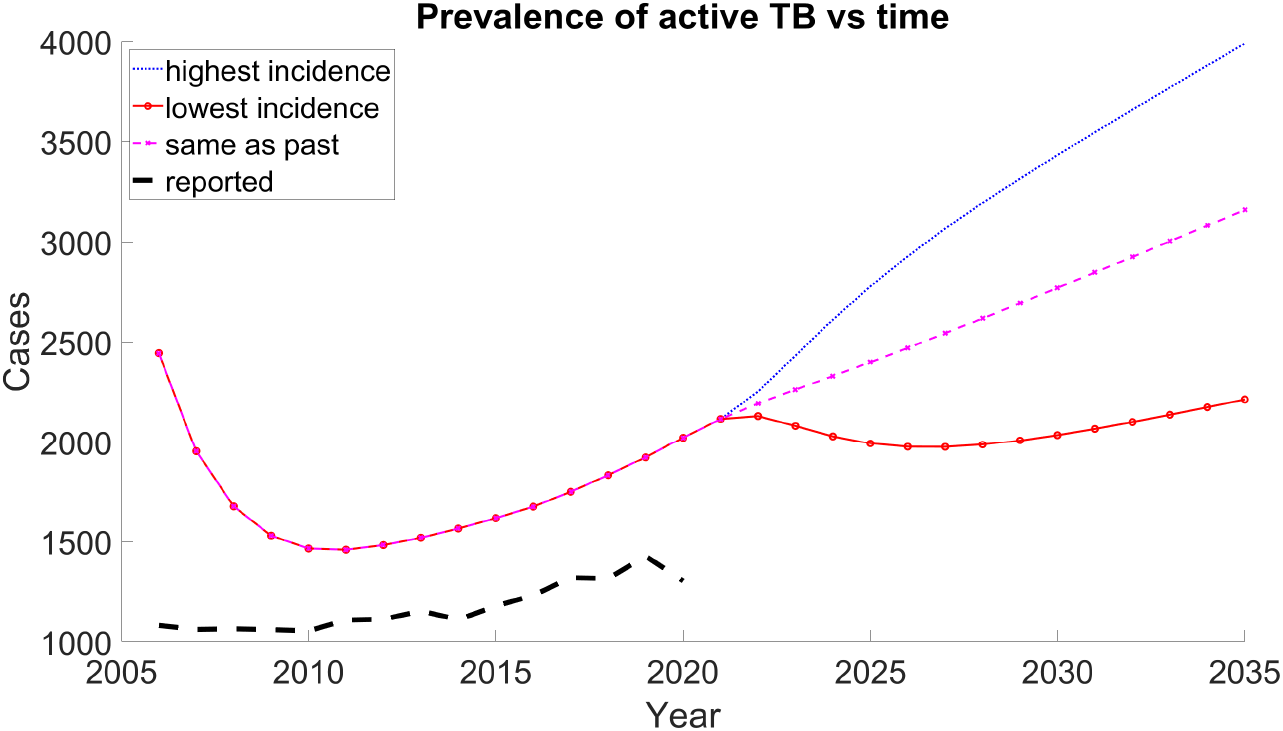
Prevalence of active TB vs time. We forecast the prevalence of active TB under different assumptions of which WHO geographic region individuals immigrate from. As in previous instances, our model estimates exceed the reported prevalence.

**Fig 13.**
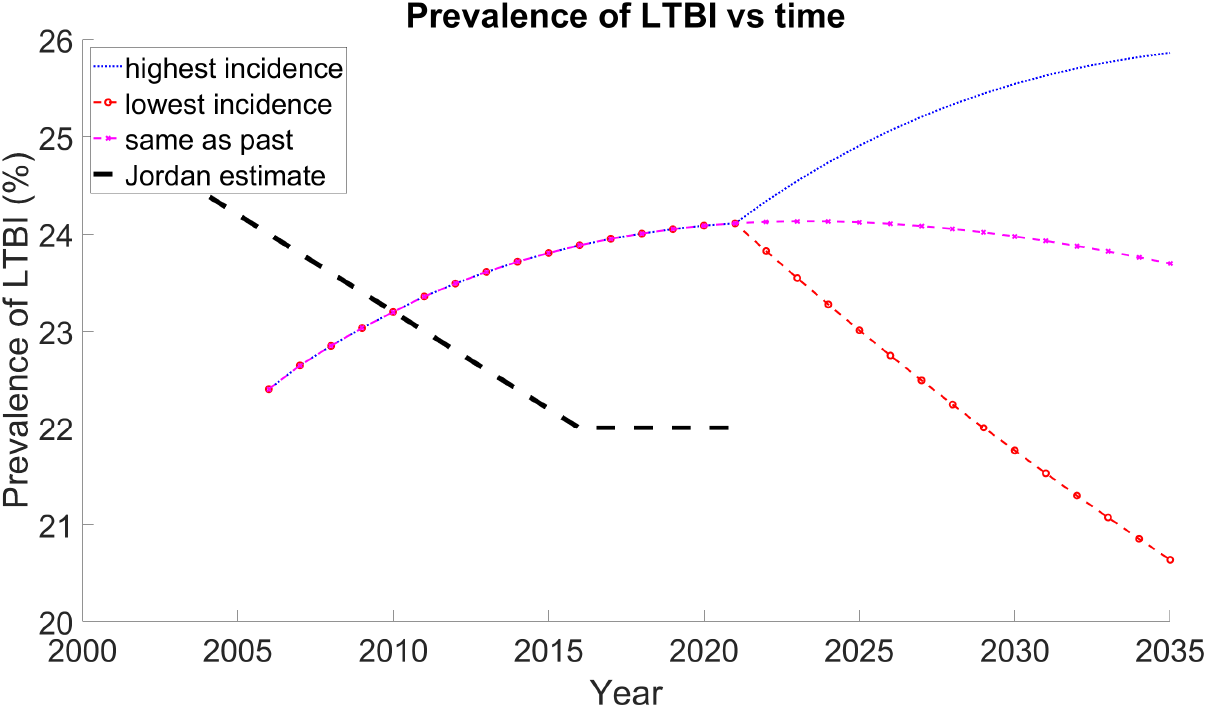
Prevalence of LTBI vs time. Although it appears our trend is reversed to Jordan’s estimate, our forecasted values are well-within the reported error margins (12).

#### Sensitivity analysis

Table 7 demonstrates the results of our sensitivity analysis, for both the incidence and prevalence of active TB in 2035. For the incidence, several parameters are reported to have total-effect index *≈* 0 because their numeric estimates were quite small (they are even sometimes negative). In fact, a study of the Monte Carlo variability in our total-effect indices (not shown) found that all effects reported as *≈* 0 are well within one standard error of zero; additional numerical estimates of the total-effect indices are presented in S4 Sensitivity analysis, Table 9. Although the parameters reported as having index *≈* 0 are small, the ranks (i.e., largest to smallest) are generally consistent across different numbers of Monte Carlo simulations.

**Table 9.** Total-effect index (incidence 2035) vs number of Monte Carlo iterations. We suspect the negative values are due to the mean index being less than the experimental standard deviation.

|  | 100,000 | 200,000 | 300,000 | 400,000 | 500,000 | 600,000 | 700,000 | 800,000 | 900,000 | 1,000,000 |
| --- | --- | --- | --- | --- | --- | --- | --- | --- | --- | --- |
| $\beta$ | -0.171425 | -0.004395 | -0.000387 | 0.006598 | 0.006343 | -0.004566 | 0.000991 | -0.000489 | 0.003110 | 0.002880 |
| $p$ | 0.057319 | 0.218293 | 0.216375 | 0.221091 | 0.225337 | 0.216167 | 0.222793 | 0.218507 | 0.222747 | 0.222165 |
| $\omega$ | -0.173688 | -0.006979 | -0.002513 | 0.004448 | 0.005504 | -0.006117 | -0.000665 | -0.002112 | 0.001705 | 0.001450 |
| $\nu$ | 0.628507 | 0.738867 | 0.746981 | 0.755689 | 0.752720 | 0.750205 | 0.750893 | 0.753431 | 0.752459 | 0.753304 |
| $n_T$ | -0.175791 | -0.006907 | -0.003216 | 0.003907 | 0.004596 | -0.006550 | -0.001150 | -0.002728 | 0.000972 | 0.000742 |
| $\delta$ | -0.176545 | -0.007503 | -0.003619 | 0.003090 | 0.004729 | -0.006808 | -0.001651 | -0.002721 | 0.000824 | 0.000601 |
| $n$ | -0.174672 | -0.006989 | -0.003359 | 0.002820 | 0.003947 | -0.006556 | -0.001793 | -0.003560 | 0.000527 | 0.000302 |
| $\sigma$ | -0.165736 | 0.015710 | 0.021724 | 0.026291 | 0.027016 | 0.016815 | 0.021306 | 0.019934 | 0.023957 | 0.023481 |

#### Incidence

We found that the incidence in 2035 was most sensitive to the parameters corresponding to activation of latent TB: the activation rate of late latent TB ν, the activation probability of early latent TB *p*, and the activation rate of recovered individuals *σ*. This is unsurprising, since the incidence of active TB *pwE* + ν*L* + *σR* explicitly depend on these parameters.

Our results show that the incidence in 2035 was not sensitive to other parameters, including the effective transmission rate *β*. This demonstrates that among the foreign-born population in Canada, the incidence of active TB is primarily driven by latently infected individuals activating rather than new infections taking place in Canada. However, another study emphasized the importance of transmission in a

#### Parameter Incidence (rank) Prevalence (rank)

low-incidence country – a genomic study conducted in Norway found that among a cohort of foreign-born immigrants with TB, about one-quarter of the patients had contracted TB in Norway (54).

#### Prevalence

For prevalence of active TB, the recovery rate δ is the most influential parameter, which is sensible since the reciprocal of the recovery rate gives the average duration an individual has active TB.

### Future work

Our model has limitations, which also motivates our future work.

#### Homogeneit

Our model assumes that the foreign-born population of Canada is homogeneous, which makes it hard to describe localized outbreaks. For instance, Colijn et al. (2007) demonstrated that in low incidence settings, local outbreaks may emerge due to heterogeneous contact structures (55). Our model also assumes the compartments are well-mixed and that everyone has equal likelihood of meeting anyone else. This could be addressed by using province-specific data (56), or by adding more compartments.

#### Fuzzy boundaries

Our model has compartments with well-defined differences between LTBI, active TB, and recovered. However, boundaries between exposed, infected, and recovered is fuzzy because transmission does not simply switch on and off (27).

#### Uncertainty quantification

While our analysis is data-driven, we do not follow the usual statistical approach of starting with a probability model for the data and developing estimators for the model parameters. In future work, we plan to extend our analysis using the Bayesian framework, incorporating prior distributions for the model parameters and giving an explicit likelihood for the data. Our model results could be used to inform the choice of prior distributions. The Bayesian approach has been successful across a wide range of epidemiological problems (57; 58; 59).

#### Reinfection

Our model assumed there was no exogenous reinfection, and that the only form of recurrent TB was due to relapse. This assumption arose because a study on recurrent tuberculosis cases in Canada and USA reported that 96% of isolates had the same genotype, and thus concluded they were due to relapse activation (35). However, Cohen et al. (2007) caution that molecular tools cannot distinguish between primary disease and reinfection (44); they also report that in moderate or low incidence settings, reinfection may be important due to contact structure and non-homogeneous mixing.

## Conclusion

To study the burden of TB among foreign-born Canadians, we developed a compartment model to describe the distribution of uninfected, actively infected, and latently infected individuals. We study the impacts of immigration by examining the number of immigrants from each source country, and the burden of LTBI there. We then validated our model by comparing to Canadian reports of TB and other model’s results. Finally, we forecast the incidence of active TB in 2035 (with assumptions about immigration and the global prevalence of LTBI) to assess whether or not Canada will meet WHO’s *The End TB Strategy* 2035 incidence goals.

Even if we assume no new infections take place in Canada, the reservoir of individuals with LTBI developing active TB will prevent Canada from meeting the End TB Strategy 2035 targets. To achieve these goals, Canada must prioritize screening and treatment of LTBI.

## Data Availability

All data produced in the present study are available upon reasonable request to the authors

## Supporting information

**S1 The burden of TB among new immigrants in 2014** *Prevalence of LTBI nationally*. In order to estimate each country’s prevalence of LTBI, we introduce some notation. Recall we earlier defined the population vector 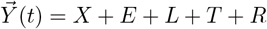, which respectively denote the susceptible, early latent, late latent, active TB, and recovered population in Canada. We similarly define this population vector for each country *c*,

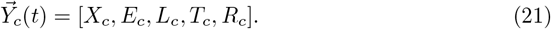

At each year *t*, the sum of *Y*_*c*_(*t*) is the total population of the country. For each country (and for each WHO geographic region), Houben and Dodd provide 3 estimates regarding LTBI in 2014:

1. the total prevalence of LTBI (denoted *HL*_*c*_),
2. the percentage of people that were recently (2 years) infected, not necessarily for the first time,
3. among those with LTBI, the percentage that were recently (2 years) infected for the first time (denoted *HE*_*c*_).

For instance, in Afghanistan (*c* = 1), estimate 1 is *HL*_1_ = 22.4%(20.8 *−* 24.1%), and estimate 3 is *HE*_1_ = 1.24%(1.02 *−* 1.45%). Notice that

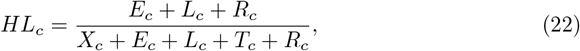

and that

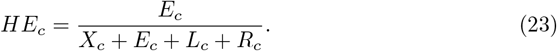

Recall that each year, the number of immigrants from each country is known and denoted *π*_*c*_(*t*). Among these *π*_*c*_ immigrants, there is a distribution of the burden of TB among new immigrants

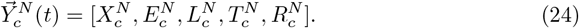

It follows that

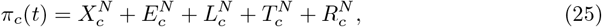

where 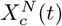 is the number of new immigrants from country *c* that have never been infected with TB. We assume that from screening, no one with active TB enters Canada, and thus for all countries *c* and years *t*

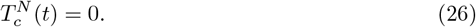

Define the total number of people worldwide in each compartment as

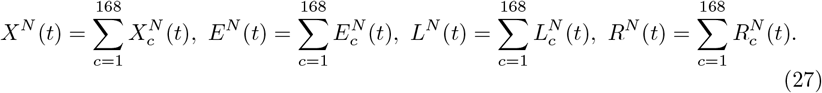

Combining Eq (25), (26), and (27) gives

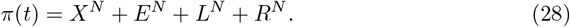

We now make two assumptions. Assume that

1. due to screening, no one with active TB enters Canada,
2. immigrants to Canada are sampled uniformly at random from among all people who do not have active TB.

Assumption 1 is reasonable and yields

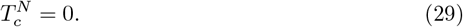

Assumption 2 is more suspect – immigration into Canada is tied to socioeconomic status of immigrants, which causes problems since burden of TB correlates with socioeconomic status. Nevertheless, with this assumption, we obtain

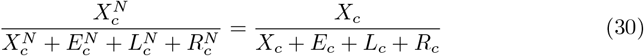

But knowing *X*_*c*_ + *E*_*c*_ + *L*_*c*_ + *R*_*c*_ is equivalent to knowing *T*_*c*_, which isn’t easy. We assume that *T*_*c*_, the prevalence of active TB, is small; for instance, even in the high-incidence country India, a metastudy from 1997-2018 estimated that the prevalence of pulmonary TB was about 295.9 per 100,000 population (60) (Canada Surveillance estimates about 70% of TB-related hospitalizations are due to pulmonary TB rather than non-pulmonary). Thus

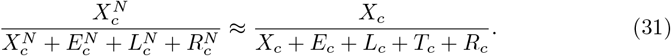

Note that the approximation Eq (31) holds if *X* in the numerator is replaced by *E, L*, or *R*.

Notice in a given year, 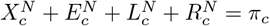. Also, denote the total population of the *c*th country at year *t* as *P*_*c*_(*t*); then *P*_*c*_ = *X*_*c*_ + *E*_*c*_ + *L*_*c*_ + *T*_*c*_ + *R*_*c*_. It follows from Eq (31) that

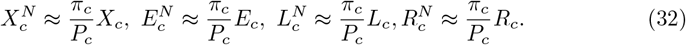

### S2 TB progression and math model details Details about the pathophysiology of TB and how it affects our math model

#### Infectiousness

Infectiousness depends on the quantity of pathogen in an infectious host, and that individual’s ability to transmit the pathogen to others. The number of bacilli excreted by persons with active pulmonary TB is relatively small, and so the probability of TB transmission per contact, per unit of time is quite low (24). The probability of transmission can be enhanced by systemic and long exposure to infectious individuals, especially in places with limited ventilation or reduced air volume per contact.

#### Recurrent TB

The proportion of recurrent TB cases due to relapse or reinfection is highly associated with the respective country (34). Cases of relapse are usually linked with drug resistance, commonly found within the first 2 years after treatment ends (61; 62). TB reinfection generally occurs in countries with high TB incidence (34), and reinfection is often linked with the coexistence of HIV (63). Having latent TB affords the hosts some protection against new infection; the amount of protection is generally hard to estimate and cannot be observed directly (64).

#### Pulmonary vs non-pulmonary TB

The PHAC reports that among hospitalizations from foreign-born Canadians due to active TB, 68% was pulmonary TB while 32% was non-pulmonary(5). Non-pulmonary TB is generally believed to be non-infectious (26). Rather than splitting our compartment *T* into pulmonary vs non-pulmonary TB, we assume the effective transmission rate *β* accounts for some individuals being non-infectious.

#### Screening for LTBI

There are two main diagnostic methods to test for LTBI: the tuberculin skin test (TST) and interferon gamma release assay (IGR). Both tests are acceptable but imperfect tests for LTBI (25). Pai et al. (2016) recommend that LTBI screening should only be performed if positive tests are supported by a serious intent to follow-up with therapy.

#### Risk factors

Pai 2016 (25) report risk factors include HIV/AIDS, type-2 diabetes, excess alcohol use, and smoking. Depression is also a risk factor(65). Narasimhan et al. (2013) additionally report malnutrition and young age are well-established risk factors to TB, and that emerging variables include diabetes, indoor air pollution, alcohol, use of immunosuppressive drugs, and tobacco smoke (66). Diedrich et al. (2020) report that HIV infection is the leading risk factor for developing severe tuberculosis (67).

### S3 Fitting unknown parameters to calibration data Details about how we fit the unknown parameters to calibration data, including the objective function

#### Fitting to reported incidence

The PHAC regularly releases reports that summarize the descriptive epidemiology of active TB in Canada (5; 46). This includes the incidence of active TB for every 100,000 foreign-born Canadians in each year (Fig 5), which we denote *f*(*t*).

We compute our model’s TB incidence at each time point, which corresponds to the rates entering the *T* compartment in Fig 3 (i.e., *pwE* + ν*L* + *σR*). We then scale so the incidence is per 100,000 individuals to obtain our model’s estimated TB incidence,

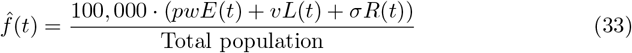

The incidence error is defined to be the squared relative difference between our computed (estimated) incidence 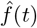 and the reported incidence *f*(*t*),

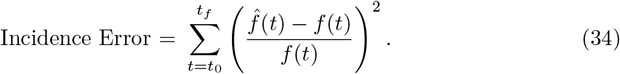

#### Fitting to the trend of reported prevalence

The PHAC reports the prevalence of active TB (46) (Fig 5). We found our model’s outputs were consistently larger than the reported data, which is unsurprising since WHO estimates as much as one-third of TB incidence is unreported (48). Hence, rather than fitting directly to the reported prevalence, we fit to the *trend* of prevalence, as described below.

Recall *T* (*t*) denotes the actual prevalence of active TB among foreign-born Canadians with active TB at time *t*. Let 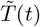 denote the reported case count(5).

We assume that in the domain *t*_0_ ≤ *t* ≤ *t*_*f*_, the reported prevalence is proportional to the actual prevalence,

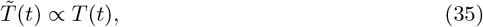

for some unknown constant of proportionality.

Define *p*(*t*) to be the ratio between the annual prevalence at year *t* and the sum of prevalence over the domain *t*_0_ *≤ t ≤ t*_*f*_ (denoted *prevalence ratio*)

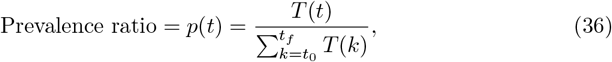

and 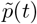 to be the corresponding reported ratio

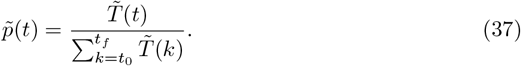

Since the proportionality constant cancels in Eq (36) and (37), we get

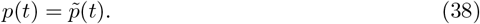

Similar to how we fit to incidence, we use our dynamic model to compute the prevalence 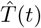,and thus determine our model’s estimate of the prevalence ratio,

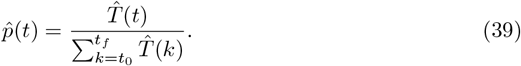

Finally, we define the Prevalence Error as

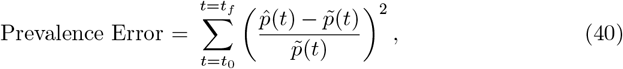

where the reported prevalence ratio 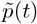 is equal to the true ratio *p*(*t*) under assumption (35).

#### Fitting relapse rate to incidence proportions

To determine *σ*, the rate at which people who have recovered from active TB may relapse and redevelop active TB, we follow the approach used by Dowdy and Cohen (2013) and Aparicio et al. (2009) (42; 43), who fit the relapse rate to the reported proportion of active TB attributed to relapse (i.e., developing active TB more than once).

Ng et al (2018) did a cohort study among 2.7 million immigrants to Canada from 2000-2013 (6). They report that 1120 of these individuals were hospitalized for active TB in Canada in 2001-2014, of which 110 of these patients had been hospitalized for active TB prior to arriving into Canada. We denote this proportion as the *proportion of incidence contributed by relapse*, or simply the *relapse proportion (RP)*, and obtain

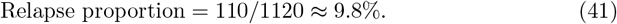

The relapse proportion obtained from our model, 9.8% (which is derived from reported data (6)), is extremely similar to the relapse proportion used by Dowdy, Dye, and Cohen (2013) (42), and approximately double the relapse proportion used by Aparicio and Castillo-Chavez (2009) (43).

*Estimating the relapse proportion*. Let 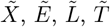, and 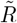 denote the number of susceptible, early latent, late latent, actively infected, and recovered individuals among the 2.7 million person cohort of Ng et al. Then the 1120 hospitalizations correspond to the total number of TB activations among latently infected individuals from 2001 to 2014,

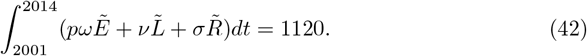

The 110 people who had active TB prior to entering Canada can be estimated in our model by the total incidence of active TB attributed to relapse,

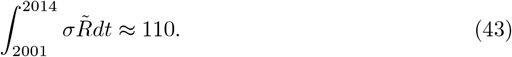

Combining Eq (42) and (43) gives an estimate of the relapse proportion,

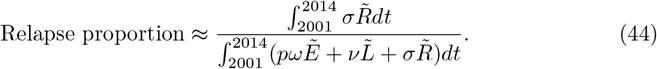

We assume that the ratio given in Eq (44) (which corresponds to the cohort of 2.7 million immigrants from (6)) is time-invariant, and that it is equal to the analogous ratio for all foreign-born Canadians. This gives our model’s estimated relapse proportion,

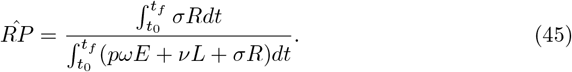

*Fitting to the relapse proportion*. We define the *Relapse Proportion Error* as

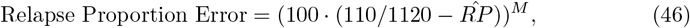

where *M* is a hyper-parameter. Notice that if *M* is a positive even integer (in practice, we took *M* = 8), the Relapse Proportion Error would be small (*<* 1) if and only if

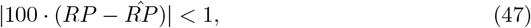

which is equivalent to

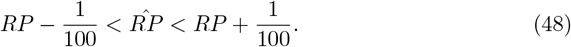

By construction, the Relapse Proportion Error calibrates the unknown parameter *σ* to ensure the relapse proportion is near the reported relapse proportion.

*Minimizing the loss function*. Optimization is performed using the `fmincon` function in `Matlab`(68). The solution vector 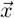 corresponds to the unknown parameters *σ q*_*E*_, *q*_*L*_, and *q*_*R*_, and the nuance parameters *X*_0_, *E*_0_, *L*_0_, *T*_0_, and *R*_0_. These values are of different orders of magnitude; hence, the data was preconditioned by scaling by a typical value (we used the initial condition as the ‘typical value’).

### S4 Sensitivity analysis

To study the sensitivity of model output to changes in parameter values, we used global sensitivity analysis methods based on the work of Saltelli (2002) and Saltelli et al. (2008) (49; 50).

Let the random variable *Y* be a function of random variables 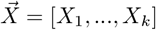,

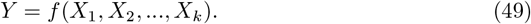

*Y* could be any model output of interest, such as the incidence and prevalence of active TB in 2035 among the foreign-born Canadian population as the output variable.

Saltelli (2002) describe the sensitivity measure for a parameter *X*_*i*_ as

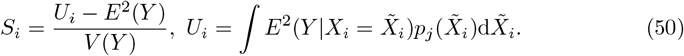

This index characterizes how much influence *X*_*i*_ has on the output *Y* by fixing *X*_*i*_ and computing the variance over all other input variables. We then average this ‘conditional variance’ over the distribution of *X*_*i*_.

The total-effect sensitivity index of a model parameter is used to show how much a parameter contributes to the variation in the model output, known as the direct effect, as well as how much a parameter’s interaction with other parameters affects the output, known as the higher-order effects. Including higher-order effects in the sensitivity index often captures the nonlinear response due to the variation in parameters.

#### Computing the indices with Monte Carlo simulation

Computing *S*_*i*_ may be inefficient, especially when dealing with a large number of parameters. Therefore, we use the Monte Carlo-based algorithm described in (49) to compute the total-effect indices.

Their algorithm requires the generation of two random input sample matrices, each of size *N* by *k*, where *N* is the number of Monte Carlo samples and *k* is the number of parameters.

#### Estimating the number of samples needed

In order to measure the sensitivity of TB incidence in 2035 on model parameters, we first run with a pilot study of 5000 experiments. This gives a mean sensitivity of 15.162 and a standard deviation of 0.614. We would like our mean to be accurate to about 3 decimal places (i.e. standard error on the order of 10^*−*3^). To this end, we use the usual formula for the standard error of a mean of independent and identically distributed random variables,

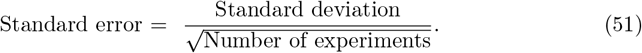

We find that a sample of size *≈* 377000 will give us a standard error of about 10^*−*3^. For safety, we use a sample size of 500000.

#### Convergence study

Recall we computed a total of 500,000 samples. To study if we expect more samples to be needed, we looked at how the total-effect indices varied as we used an increasing number of samples.

We computed the total-effect indices using 5000 randomly selected samples (repeating a total of 10 times and averaging the results). We then repeated for 54500, 10400… 450000. The average total-effect indices are displayed in Fig 14. Notice that as the number of iterations increases, the total-effect indices stabilize.

**Fig 14.**
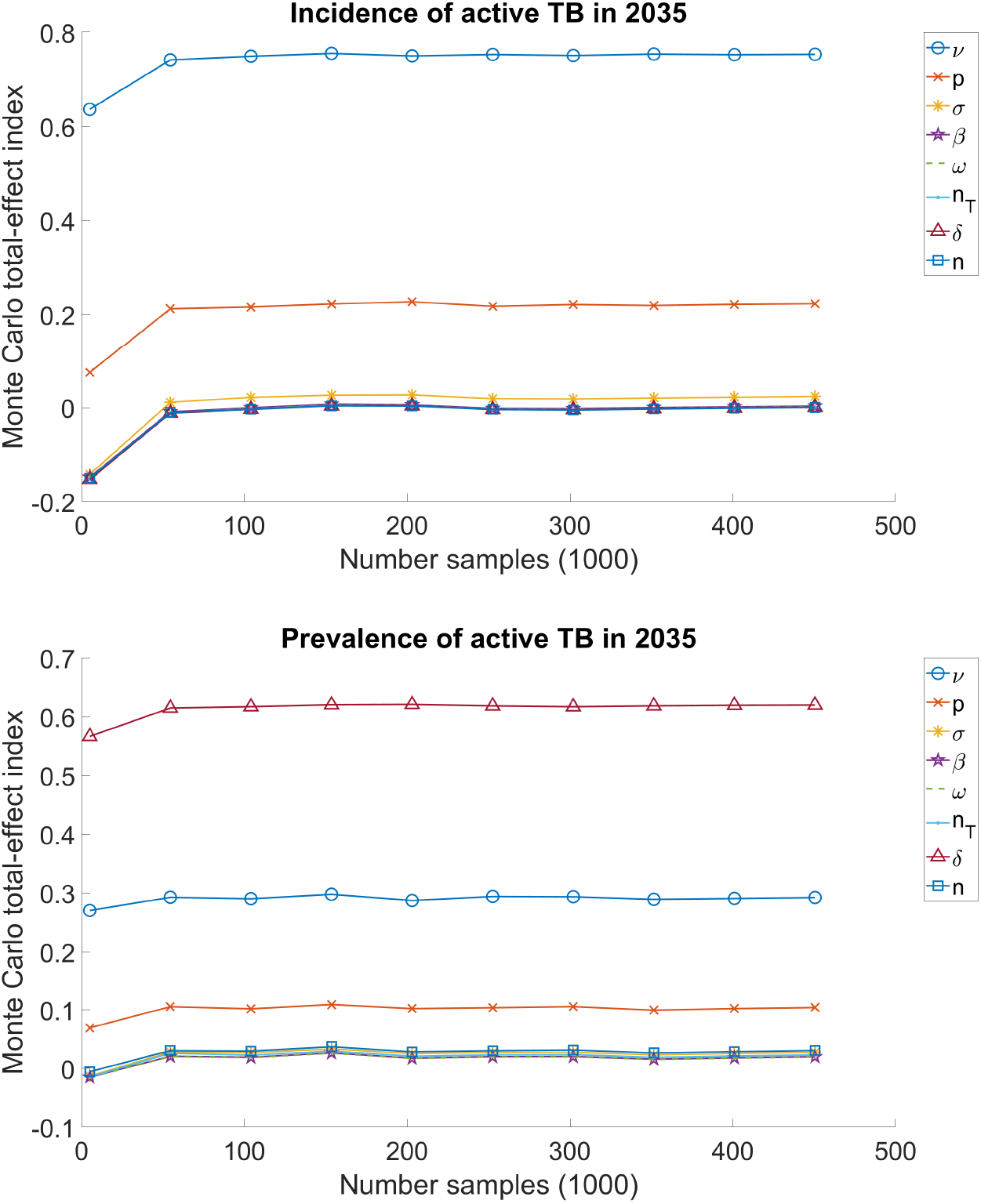
Total-effect index vs total number of Monte Carlo samples. Incidence of active TB is most affected by the activation rates of LTBI to active TB, ν, *p*, and *σ*. Prevalence of active TB is affected most by the recovery rate δ, and then by ν and *p*.

#### Generating random samples

To generate one random sample, *k* random parameters must be created. We generate each parameter from a uniform distribution with centers matching the plausible values in our Set 1 from Table 5 and endpoints chosen to be *±*10% from the center.

For instance, the random variable *X*_6_ (which corresponds to the recovery rate δ) is drawn from *U*(*a*_6_, *b*_6_), the uniform distribution on the interval (*a*_6_, *b*_6_), where

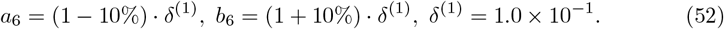

We similarly construct intervals for the remaining parameters, *X*_1_ = *β*(1), …, *X*_8_ = *σ*^(1)^.

#### Visual sensitivity analysis

In this section, we describe our treatment of the estimated and unestimated parameters in more detail, and discuss what outputs we extract from our individual simulations and sensitivity analysis.

Those parameters not estimated are fixed at levels determined from the medical and epidemiological literature. In order to explore the impact of these parameters on the overall model, we select a range of values for each and repeat the analysis described in *Specifying Parameters* at every combination of parameter values. This gives us a measure of how sensitive our findings are to each parameter over the range of values we explore. For more on sensitivity analysis, see Razavi et al (69).

Each non-estimated parameter has three levels (see Tables 2), and there are a total of 3^8^ = 6561 combinations. For each such parameter, we divide all analyses based on the levels of that parameter. We then plot all incidence trajectories obtained from a single level of that parameter in the same plot, along with the average trajectory. We then plot the average trajectory for each level of the parameter, as well as the global average trajectory. This combination of plots gives a visual representation of the relative variability of estimated incidence across levels of the parameter. Comparing plots across parameters allows us to assess the relative sensitivity of our model to each non-estimated parameter.

Fig 15 displays a summary of all parameters explored, Fig 16 demonstrates the sensitivity of early activation probability *p* to incidence, and Fig 17 highlights the insensitivity of infectivity *β* to incidence.

**Fig 15.**
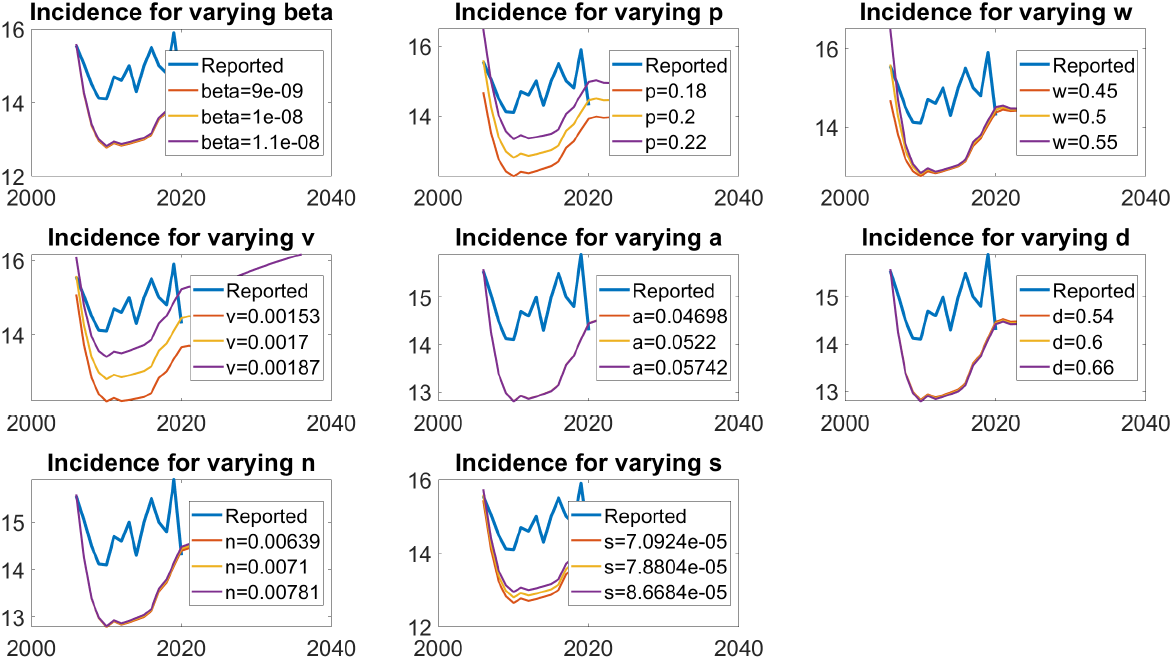
We can visually see that incidence is most sensitive to parameters *p*, ν, and *σ*.

**Fig 16.**
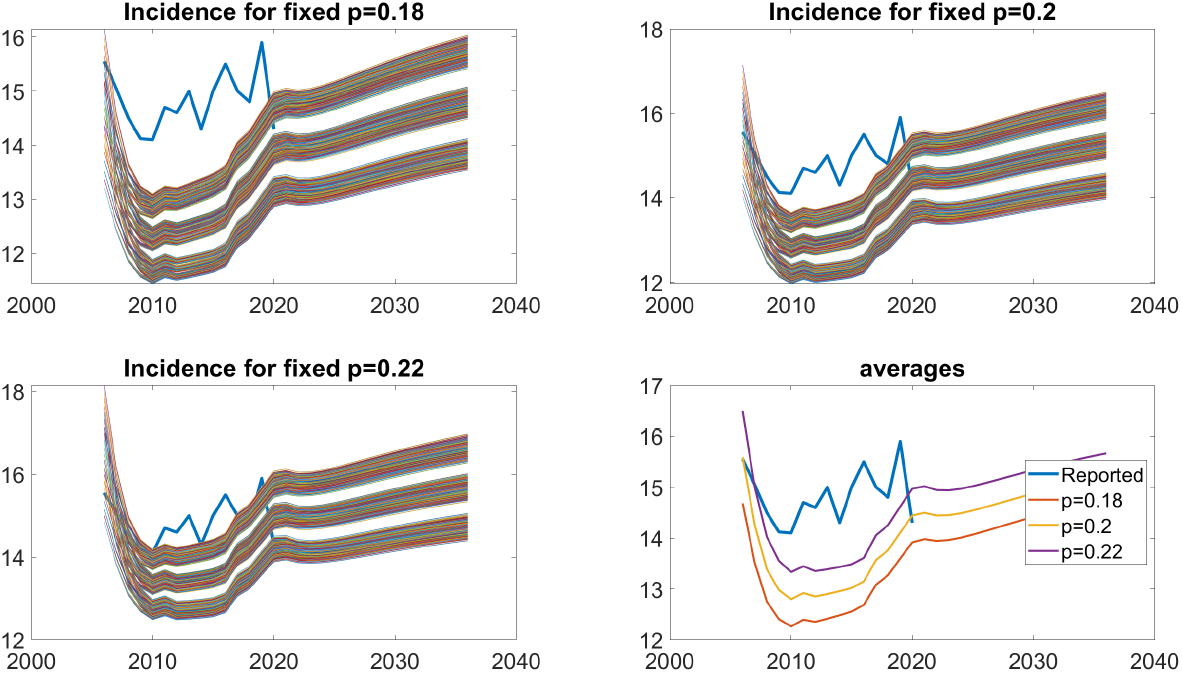
Sensitivity of early activation probability *p* on incidence. The first 3 subfigures demonstrated simulated incidence trajectories for a fixed value of *p*. In the last subfigure, each trajectory corresponds to the average value of all trajectories for a fixed value of *p*.

**Fig 17.**
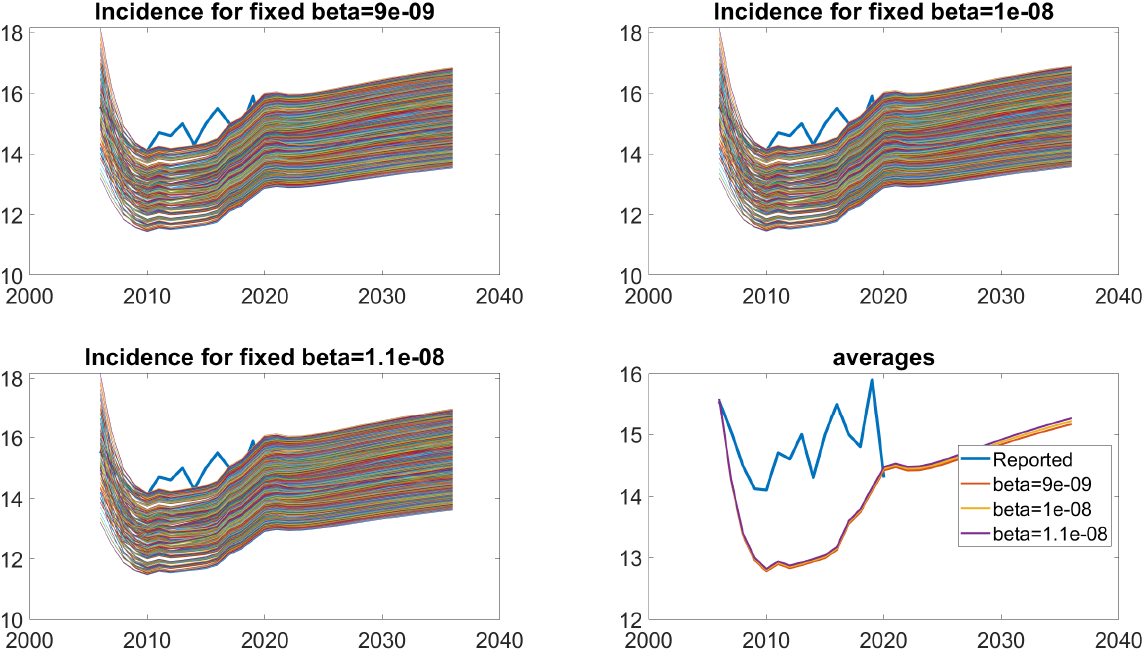
Sensitivity of infectivity *β* on incidence. These trajectories are not unified, suggesting the incidence is not sensitive to infectivity *β*.

### S5 Forecasting the burden of LTBI among new immigrants

The burden of TB among new immigrants is given by *q*_*X*_, *q*_*E*_, *q*_*L*_, *q*_*T*_, and *q*_*R*_. We first determine these values in the year 2014, then make assumptions to estimate them in previous and subsequent years. Again, we again assume *q*_*T*_ = 0 due to screening.

Following the notation used in S1 The burden of TB among new immigrants in 2014, recall that *X*^*N*^ (*t*) is the population of susceptibles among new immigrants entering Canada in year *t*, and *π*(*t*) is the number of new immigrants entering Canada in year *t*. Notice that

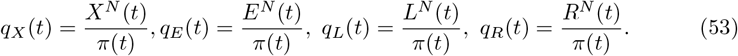

Additionally, the number of new susceptibles is simply the sum across the corresponding number from each WHO geographic region,

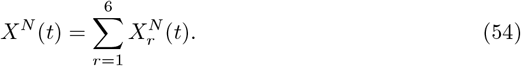

Thus, to compute *q*_*X*_, *q*_*E*_, *q*_*L*_, and *q*_*R*_ in 2014, we need 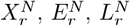, and 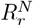 for every WHO geographic region in 2014, which requires four equations. They are the following.

1. *Annual new immigrants by WHO geographic region*. By definition, *π*_*r*_(*t*) immigrants entered Canada from region *r* at year *t*, so

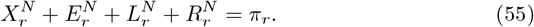 We assume that *π*_*r*_(*t*) is known.
2. *Houben’s estimate of LTBI prevalence*. This combines Eq (31) and (22). Let *HL*_*r*_ denote Houben’s estimate of the prevalence of LTBI in country *r* in year 2014. Then

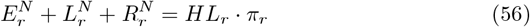 Ranges of *HL* and *HE* for each WHO region is reported in Table 1.
3. *Houben’s estimate of new latent TB*. This combines Eq (31) and (23). Let *HE*_*r*_ denote Houben’s estimate of the percentage of people in region *r* with LTBI who were recently (2 years) infected. Then

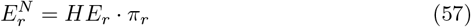
4. *Empirical ratio of q*_*L*_ *to q*_*R*_. Of the 4 experiments that passed our feasibility study (Table 5), we found 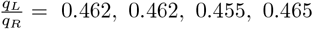. So we assume *q*_*L*_*/q*_*R*_ = 0.46. We naively apply this to every region, so that

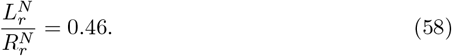

Finally, computing *q*_*X*_, *q*_*E*_, *q*_*L*_, *q*_*R*_ from Eq (53) in 2014 gave

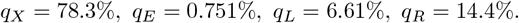

These results are similar to optimizer outputs, Table 5.

*Extending beyond 2014*. We again assume that prevalence decays exponentially. Following the methodology presented in Eq (18), we have the prevalence of LTBI in WHO geographic region *r* at time *t*

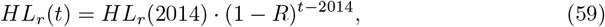

where *R* = 1%.

On the other hand, we assume that *HE*_*r*_ remains constant in time.

## Acknowledgments

We received funding from Langara’s Work on Campus program, Student Work Assistance Program, and Applied Research Centre; and from SFU’s Dean of Graduate Studies. We would also like to thank SFU’s MAGPIE group, especially Caroline Colijn’s TB journal club, for the informative discussions; James Park for his literature review; Dominic Lam for his numerical scheme to solve our differential equations; Alexandra Wong, Tiffany Yu, and Ailene Macpherson for their suggestions on our figures; Alexander Beams and Haniel Yan for their valuable feedback; and Katie Wong for minding many of my *Mycobacterium tuberculosis* monologues.

William Ruth was supported by a CANSSI postdoctoral fellowship.

